# Nasal Gene Expression in ART-Naive Adults with HIV and Pulmonary Tuberculosis in Uganda

**DOI:** 10.64898/2026.01.06.26343354

**Authors:** Nisreen Khambati, Kattya Lopez Tamara, Elizabeth Nakabugo, Arthur Van Valkenburg, Jessica K Anderson, Sean Lu, Rinn Song, Suryaram Gummuluru, Andrew J Pollard, Jerrold Ellner, Padmini Salgame, Else Margreet Bijker, Lydia Nakiyingi, Daniel O Connor, W Evan Johnson

## Abstract

**Background:** Diagnosis of pulmonary tuberculosis (TB) in people living with HIV remains difficult. Since the first pathogen-host interaction in TB occurs in the upper airway, we hypothesized that host transcriptomic analysis on nasal specimens may identify novel diagnostic biomarkers. We aimed to demonstrate differences in nasal gene expression between people with HIV and TB disease versus people with HIV without TB, evaluate the performance of nasal signatures in classifying TB and compare nasal gene profiles with blood gene profiles from the same cohort.

**Methods:** We enrolled adults in Uganda with newly diagnosed HIV and symptoms of pulmonary TB disease We collected nasal cells and blood for RNA sequencing to identify differentially expressed genes and enriched pathways between people with HIV and TB disease and people with HIV without TB. Supervised machine-learning of gene expression data was used to predict TB status.

**Results:** 40 adults living with HIV were enrolled (median age: 34 years, median CD4 count: 182), including 20 with TB disease and 20 without. We identified 44 nasal differentially expressed genes and 238 blood differentially expressed genes, with three overlapping genes between sample types. Models trained using all 44 nasal differentially expressed genes had a cross-validated area under the curve between 0.87-0.90 for predicting TB disease amongst adults living with HIV. A simplified four-gene signature (*SPIB, SHISA2, TESPA1* and *CD1B*) met the World Health Organization criteria for a TB triage test. Among adults with TB, pathways related to the inflammatory response and innate immune system were downregulated in nasal samples and upregulated in blood.

**Conclusion:** This proof-of-concept study demonstrated that there were distinct nasal gene expression patterns associated with TB, not seen in blood. Differences in nasal gene expression in people with HIV who have TB disease, versus those without TB, highlight their potential as diagnostic biomarkers. Further validation studies of gene signatures using minimally invasive nasal samples are recommended in other difficult to diagnose groups.

## Background

Tuberculosis (TB) is the primary reason for hospitalization and death in people living with HIV (PLHIV)^1^, with the highest TB-HIV coinfection in the African Region^2^. TB-related mortality in PLHIV is partly due to difficulties in diagnosis. Autopsy studies in PLHIV demonstrated that TB was undiagnosed in nearly half of cases identified post-mortem^3^. PLHIV are more likely to have paucibacillary disease and difficulty in expectorating sputum than those without HIV^1^, limiting sputum-based tests such as culture or Xpert MTB/RIF Ultra (Ultra), a cartridge-based polymerase-chain-reaction (PCR) assay. Recognizing this, the World Health Organization (WHO) has advocated for development of non-sputum-based biomarker tests for diagnosis of pulmonary TB^4^.

Several blood-based mRNA biomarkers have shown promise as triage tests for TB^5^. However, their potential has been limited by a poorer performance in PLHIV^6,7^, particularly in antiretroviral therapy (ART) naive individuals^8^, or when differentiating TB from other respiratory diseases^9^. Lower specificity in PLHIV could be due to the dominance of interferon-stimulated genes (ISGs) in blood and HIV-related viraemia or opportunistic infections upregulating interferon-driven pathways^8-10^. Therefore, alternative sample types for biomarker discovery are worth exploring. With the first pathogen-host interaction in TB occurring in the upper airway, transcriptomic analysis of nasal specimens may identify host gene profiles and pathways not previously identified in blood. Overlap in patterns of gene expression across the upper and lower airways in respiratory inflammatory conditions^11-13^ suggests that nasal samples could be proxies for studying lung disease. Moreover, nasal specimens are also minimally invasive and easy to collect.

A rich transcriptomic response in nasal samples has been demonstrated for biomarker discovery and understanding pathophysiology for several non-TB respiratory illnesses, including respiratory syncytial virus^14,15^, SARS-CoV-2^16-18^, pneumonia^19^, asthma^20^ and lung cancer^12^. Pandya *et al* analyzed 1,555 nasal samples across ten cohorts and developed a 33-gene nasal signature that could classify viral acute respiratory infections from healthy or non-viral samples with⍰over⍰80% specificity and sensitivity^21^. Amongst SARS-CoV-2 patients, suppressed interferon responses in nasal samples were linked to more severe disease^17^. In children, nasal gene signatures could accurately predict respiratory viral infections^14,22^, and in some cases, performed better than blood classifiers^14^. Such studies highlight the value of studying the local nasal response to respiratory infections.

Despite these potential benefits, to our knowledge, nasal gene expression profiling has not been studied in TB. To address this gap, we sought to determine if nasal gene expression differs in ART-naive PLHIV and pulmonary TB disease compared with PLHIV without TB disease. We identified differentially expressed genes (DEGs) and biological pathways from nasal cells and assessed the performance of nasal signatures in classifying TB disease. As secondary objectives, we compared nasal-derived DEGs, biological pathways and gene signatures with paired blood samples from the same cohort and evaluated the diagnostic performance of existing published blood TB signatures in our nasal dataset.

## Methods

Figure 1 provides an overview of the methods. Computational code for the methodology is available at: https://github.com/nisreenkhambati/uganda_nasal_cells.

**Figure 1.**
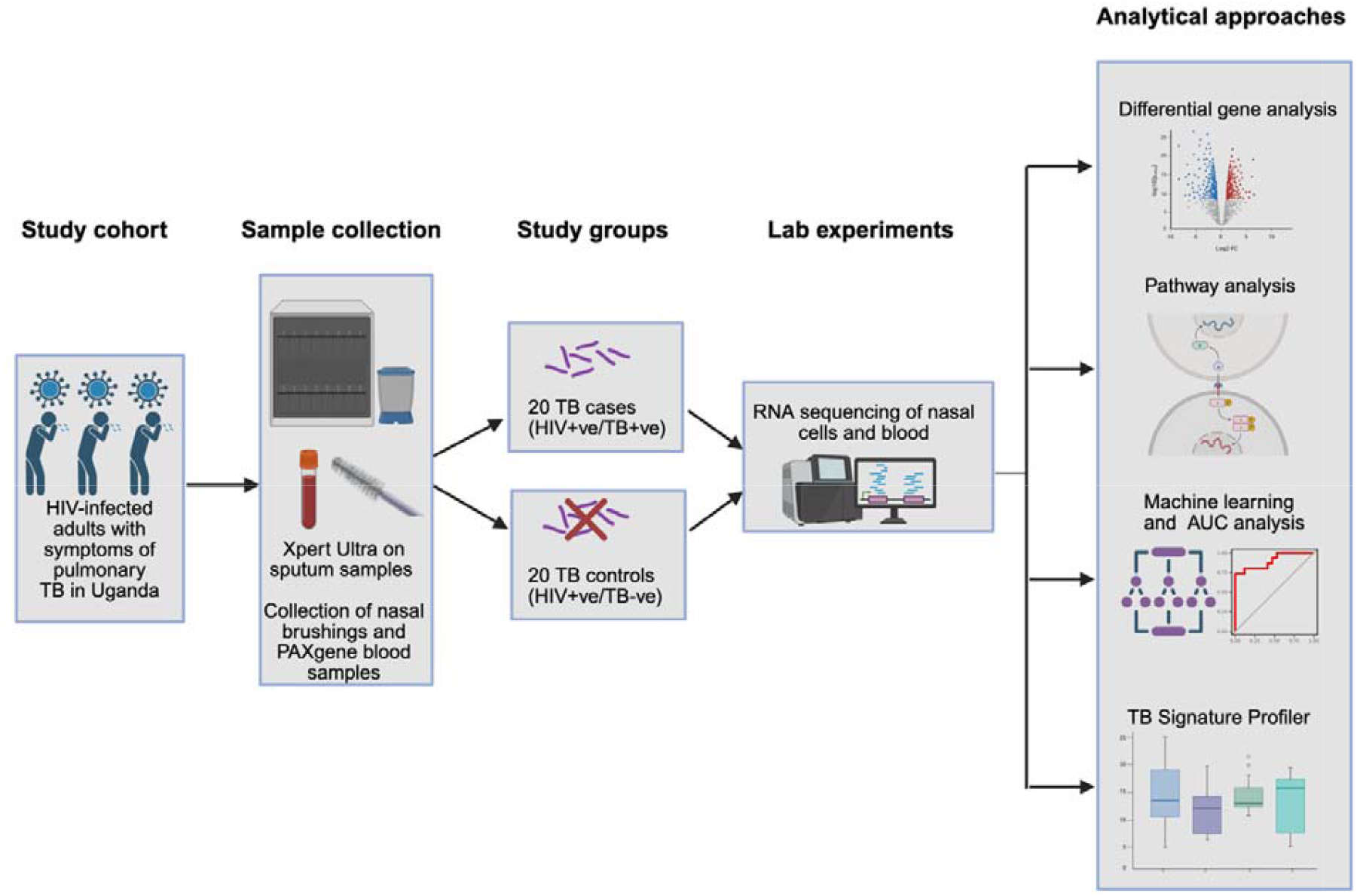
Overview of the methods. Adults in Uganda with newly diagnosed HIV and TB symptoms were enrolled and nasal and blood samples were collected for RNA sequencing. Analysis aimed to determine if nasal gene expression differed in PLHIV and pulmonary TB disease compared with PLHIV without TB disease. Figure created in BioRender.com, license obtained by Anderson, J., 2026, https://BioRender.com/3fuig8f) Ultra= Xpert MTB/RIF Ultra; AUC= area under the curve; PLHIV=people with HIV

### Study cohort

This study was conducted at Mulago National Referral Hospital and Infectious Diseases Institute-affiliated clinics in Kampala, Uganda between 2020 and 2021. Individuals between 18-50 years with presumptive TB (defined as having symptoms of pulmonary TB including cough, fever, night sweats, weight loss and hemoptysis), no previous HIV or TB treatment (including isoniazid prophylaxis) and newly confirmed HIV were consecutively enrolled. The HIV status of those with presumptive TB was determined as part of study screening, i.e. HIV was diagnosed due to the presentation with TB symptoms. Those who were current or former smokers, had diabetes, severe malnutrition, a history of TB, or had used corticosteroids or antibiotics in the past three months were excluded. Informed consent was obtained from all participants, and the study was approved by institutional review boards of all partnering institutions (Boston University Medical Campus, Joint Clinical Research Centre and Uganda National Council for Science and Technology).

### Definitions and sample collection

Each adult had one early morning expectorated sputum tested with Ultra (Cepheid, CA, USA). Adults were classified as having active TB disease based on a positive sputum Ultra. Control subjects were those with no microbiological evidence of TB.

Study personnel followed a standardized protocol to collect nasal cells and blood. A cytology brush was used to collect nasal cells by insertion into each nostril and rotating it against the lateral wall for three seconds. The brush head was placed into a cryovial containing RNA-preserving agent. Blood was collected into PAXgene RNA tubes. Samples were obtained before TB or HIV treatments were initiated and frozen at -80°C.

### RNA sequencing and quality control

Nasal and blood samples were shipped on dry ice to Rutgers University. Total cellular RNA from nasal and PAXgene blood samples was extracted using the RNeasy Plus Micro Kit protocol (QIAGEN Inc., Hilden, Germany) at Rutgers University according to the manufacturer’s instructions. The integrity of RNA was analyzed using an Agilent TapeStation. To generate the cDNA library, the NEBNext® rRNA Depletion Kit was used to remove the ribosomal RNA. Illumina-compatible RNA-seq library was then prepared using the NEB Next Ultra II RNASeq library preparation kit. The cDNA libraries were purified with AmpureXP beads and quantified using an Agilent TapeStation and a Qubit 4 Fluorometer. Equimolar amounts of barcoded libraries were pooled and sequenced on the Illumina NovaSeq S2 FC 6000 (Illumina, San Diego, CA), producing 40-50 million paired-end 150 base-pair reads per sample. All samples were run across two lanes in a single flow cell in the same sequencing run.

Raw FASTQ data were assessed for quality using FastQC^23^. Trimmomatic was used to remove low quality bases and adaptors (LEADING:3 TRAILING:3 SLIDINGWINDOW:4:15 MINLEN:36)^24^. Reads were aligned to the human genome (GRCh38.p14) using the Spliced Transcripts Alignment to a Reference (STAR) tool^25^ and annotated using the GENCODE v44 primary assembly.^26^

Gene count data were exported to R (version 4.3.1) and subset for protein coding genes using the biomaRt package^27^. Mitochondrial, hemoglobin, ribosomal and RNA genes, as well as low-count genes, were filtered out. Outliers were identified by examining the Euclidean distances between pairwise combinations of samples, visualized using ComplexHeatmap^28^ and principal component analysis (PCA). TB status, age, sex, CD4 count, and HIV stage were investigated as potential expression-associated variables using PCA and corrected if needed (excluding TB status) for downstream analyses. Differences in principal components for variables separating TB and controls were statistically tested using the Student’s t-test, given normality of data distribution. All PCA and heatmap analyses were performed on transformed normalized counts (variance stabilizing transformation).

### Differential gene expression, machine learning and ROC analysis

Differential gene expression analysis between PLHIV and TB disease and PLHIV without TB disease (controls) was performed in the nasal and blood datasets using DESeq2^29^. DEGs were defined as having an adjusted p-value of <0.05 and log2 fold-change ≥1. Identified DEGs were used to cluster samples, visualized with ComplexHeatmap^28^, using Euclidean distance as a measure of dissimilarity and complete linkage for between-cluster separation. Gene set enrichment analysis (GSEA) was performed using fgsea^30^ to evaluate whether DEGs from one tissue were enriched in the ranked list of genes from the other tissue. Genes were ranked based on the test statistic from DESeq2^29^.

Models were trained with six machine learning algorithms based on all DEGs using caret^31^: random forest (RF), support vector machine (SVM) with radial and linear kernels, elastic net, partial least squares (PLS) discriminant analysis and k-nearest neighbors (KNN), with 10-fold cross-validation repeated 10 times. These models incorporate both linear and non-linear data relationships and are commonly used in TB diagnostic transcriptomic research^32-36^. Prior to model training, gene counts of nasal and blood DEGs underwent variance stabilizing transformation. Predictors were pre-processed by removing near zero-variance variables and centering and scaling for models sensitive to feature scaling (elastic net, SVM, KNN and PLS) were also performed. Receiver operating characteristic (ROC) curves for each model in diagnosing TB were constructed with the MLeval package^37^ to estimate the area under the curve (AUC), sensitivity, and specificity with 95% confidence intervals. Cut-off thresholds were determined by the maximum Youden index^38^.

Models with a reduced gene panel are potentially easier to translate to a diagnostic test. Variable importance for each nasal model trained using all DEGs was estimated using caret^31^. The top 10 predictive genes for each model were identified and genes common to all six nasal models were retained to refit a new model using only these genes. The AUC, sensitivity and specificity of this parsimonious nasal signature in classifying TB was calculated using MLeval^37^.

Finally, the INTERFEROME database^39^ was used to identify which top DEGs were interferon-stimulated genes, which we defined as genes that were significantly up or down regulated (>2.0-fold change, p<0.05) in human samples treated with interferon versus controls.

### Pathway analysis and sample deconvolution

Pathway analyses for nasal and blood samples were performed with GSEA using the fgsea package^30^, and over-representation analysis (ORA) using the clusterProfiler package^40^. For fGSEA, ranked gene lists using the test statistic were compared to Hallmark gene sets and KEGG and Reactome pathways from the Molecular Signatures Database (MSigDB)^41^. For ORA, gene ontology biological processes^42^ (GO BP) were evaluated in the differentially expressed upregulated and downregulated genes separately in adults with TB. Pathways with adjusted p-value < 0.05 were considered functionally enriched.

Nasal and blood gene lists from DESeq2 were also uploaded to the Qiagen’s Ingenuity Pathway Analysis (IPA) tool^43^ and filtered by adjusted p-value (false discovery rate) < 0.1 to determine canonical pathways and upstream regulators in TB. IPA uses a Fisher’s exact test to compute p-values and uses the log2 fold-change to compute z-scores. For visualization, canonical pathways and upstream regulators with a p-value <0.05 and IPA z-score≥ ±1.5 were shown.

Nasal samples consist of both immune and epithelial cell components. The CIBERSORTX tool and the validated LM22 gene signature matrix^44,45^ were used to examine the immune cell composition in nasal and blood samples. CIBERSORTX is a deconvolution tool that employs support vector regression to estimate the relative proportion of immune cell subtypes from mixed bulk tissue transcriptome profiles^45^. The LM22 matrix consists of 547 genes that can accurately distinguish 22 immune cell populations, which can further be summarized into 11 major cell types based on shared lineage, including B cells, CD4 cells, CD8 cells, gamma delta T cells, eosinophils, plasma cells, NK cells, macrophages, neutrophils, dendritic cells and mast cells^44^. Counts data were first normalized as transcripts per million (TPM) to match the signature matrix. CIBERSORTX was performed with B-mode batch correction, quantile normalization disabled, and the number of permutations set to 500. Results were expressed as fractions of total leukocyte content. Proportions of estimated immune cell populations were compared by TB status using either Student’s t-test or Mann-Whitney U test depending on normality of data distribution, with analyses corrected for multiple comparisons.

### Performance of published blood RNA signatures

The TBSignatureProfiler package^46^ was used to apply published blood signatures that diagnose TB disease to our nasal and blood RNA sequencing datasets. We selected blood signatures that were shown to distinguish TB disease from latent TB infection, TB from some combination of other diseases or TB from HIV. Raw counts were first converted into log counts per million (CPM). Signature performance scores were calculated using gene set variation analysis (GSVA) and single-sample GSEA (ssGSEA) and compared using two-sample t-tests and bootstrapped AUCs. Only published blood signatures with two or more genes that coincided with genes in our sequencing data were used in the analysis.

## Results

### Study participants

Forty PLHIV were enrolled, including 20 with pulmonary TB disease and 20 without (controls). The median age was 35 years, 45% were male, 55% had HIV stage 3 and the median CD4 count was 182. PAXgene samples were collected from all, whereas nasal samples were available from 35 (17 with TB and 18 controls). Adults with TB and controls did not differ in age, sex, CD4 count or HIV stage **(Supplementary Table 1)**.

### Quality control and PCA

67% of nasal and 57% of blood reads uniquely mapped to the human genome **(Supplementary Table 2)**. Three nasal samples (S4, S20, S31) demonstrated high average sample-to-sample distances from most other samples and had low reads (<5 million reads) **(Supplementary Figure 1)**. These samples were removed, with 15 adults with TB and 17 controls remaining for downstream nasal analyses. No blood samples failed quality control, hence none were excluded.

Using PCA, nasal samples from adults with TB and controls clustered differently on PC4 (p= 0.048) **(Supplementary Figure 2A, 2C)**. In blood, differences in PC1 between TB and controls were significant (p = 6.853×10^−4^) **(Supplementary Figure 2B, 2D)**. Males clustered separately from females for PC3 (p= 2.798 ×10^−3^) and PC4 (p= 4.943 ×10^−7^) in nasal samples and for PC3 (p=5.449 ×10^−5^) and PC4 (p=1.303 ×10^−5^) for blood, whereas HIV-related variables and age did not separate samples **(Supplementary Figure 3)**. After sex-correction, PC3 in nasal samples and PC1 in blood samples demonstrated significant differences between TB and controls (nasal PC3: p = 0.046, blood PC1: p = 2.706 ×10^−4^) **(Supplementary Figure 4)**.

### Differential gene expression and ROC curve analysis

44 nasal DEGs and 238 blood DEGs were identified between TB and controls **(Figure 2, Supplementary Table 3)**. Nine of the top ten blood DEGs were classified as ISGs compared to four of the top nasal DEGs. Hierarchical clustering based on the DEGs showed grouping of samples by TB status, although overlap remained between the two groups **(Supplementary Figure 5)**.

**Figure 2.**
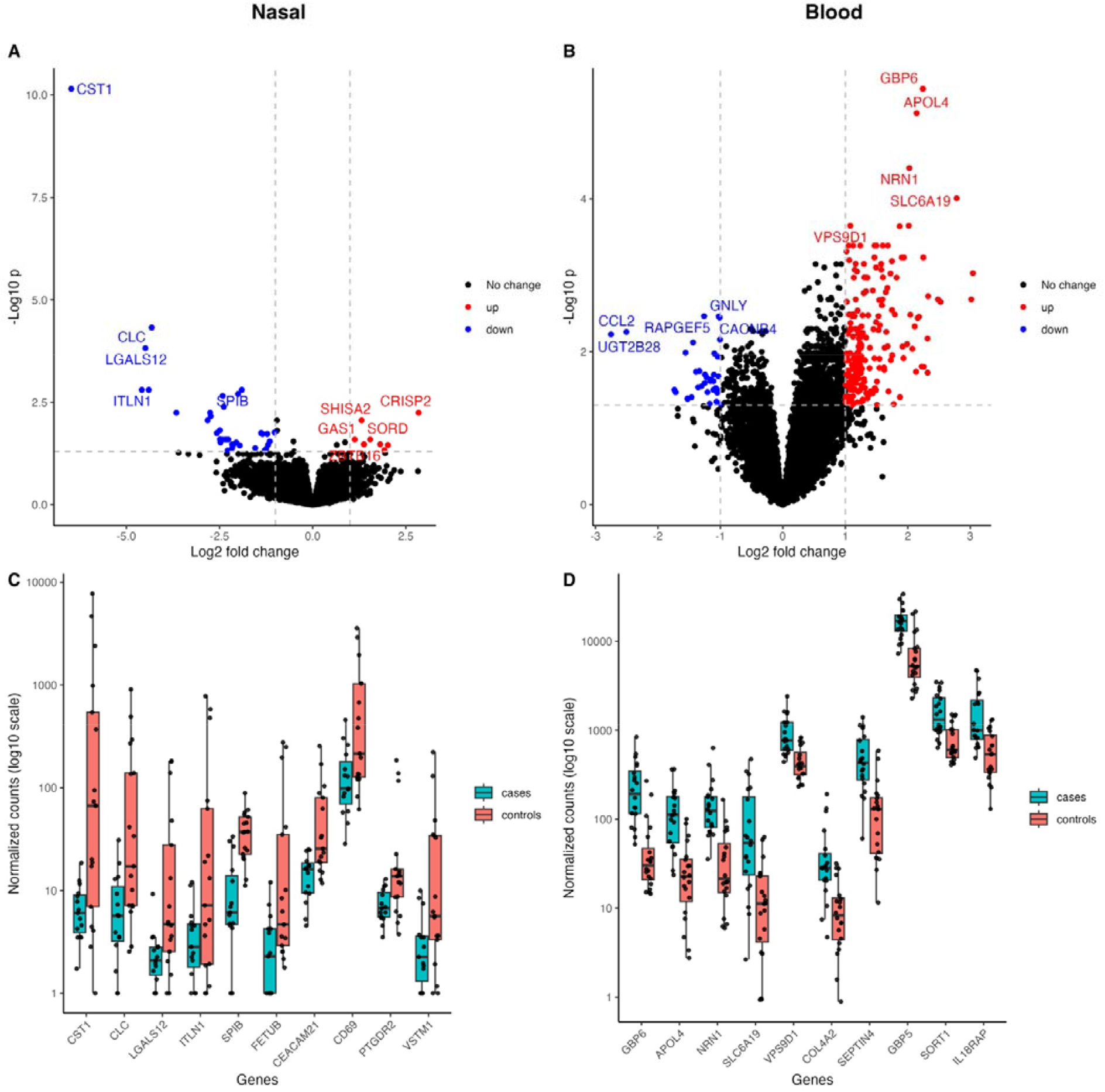
Volcano plots and box plots illustrating the top DEGs in PLHIV and TB compared to PLHIV without TB (controls). Volcano plots show the effect size by the -log_10_P value for nasal (A) and blood (B) samples. DEGs are highlighted in red (upregulated) or blue (downregulated), with gene names shown for the top 5 upregulated and downregulated DEGs. Boxplots are of normalized counts (log_10_ scale) for the top ten DEGs, which were all downregulated for nasal samples (C) and upregulated for blood samples (D) in TB cases. Individual data points are shown as jittered dots. DEGs=differentially expressed genes

In TB compared with controls, most nasal DEGs were downregulated whereas most blood DEGS were upregulated, with only three overlapping DEGs **(Figure 3)**. Based on fGSEA, nasal DEGs were significantly negatively enriched in the ranked blood gene list and blood DEGs were also negatively enriched in the ranked nasal gene list **(Supplementary Figure 6)**, suggesting that genes respond differently to TB disease in the two tissues, with compartment-specific immune responses.

**Figure 3.**
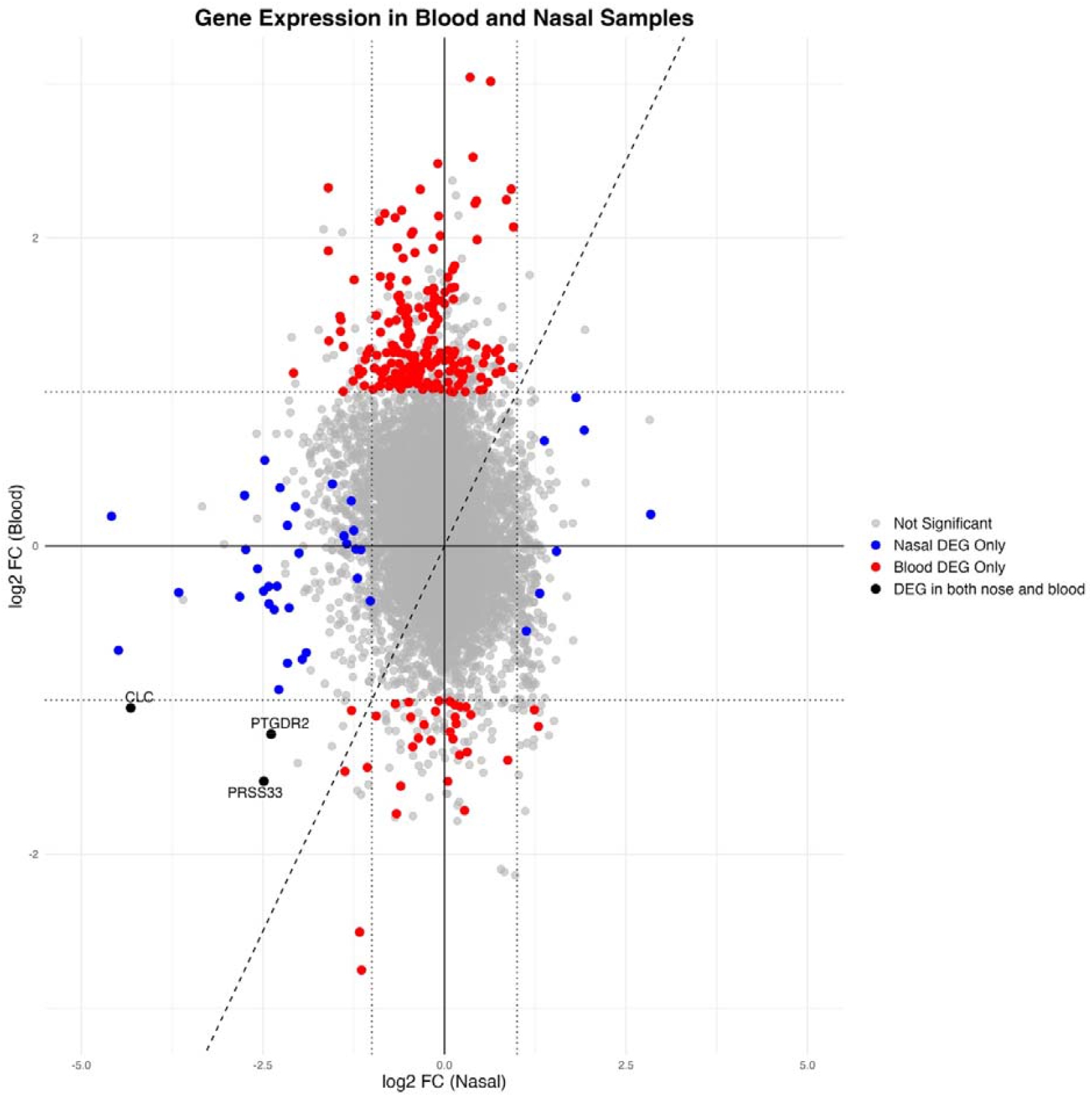
Agreement plot of log2FC for genes in blood samples (x-axis) and nasal samples (y-axis). Colored features are genes which were differentially expressed with log2 FC ≥1 and an adjusted p-value of <0.05. Dashed x=y line represents agreement. Dotted lines are shown at +/-1 log2FC. Labelled DEGs are those which are common to both nasal cells and blood and were downregulated in TB. DEGs= differentially expressed genes; log2 FC= log2 fold-change.

Models trained using all 44 nasal DEGs had a cross-validated AUC between 0.87-0.90 for predicting TB status in the nasal dataset, which was similar to models trained with the 238 blood DEGs (AUC: 0.89-0.98) in blood (Figure 4, Table 1). The highest AUC for the nasal models was RF. All four adults with TB that were missed by the nasal RF model were correctly identified as TB by the blood RF model. Likewise, the two adults with TB missed by the blood RF model were correctly classified by the nasal RF model.

**Table 1:** Diagnostic performance with 95% CI of each model.

|  | NASAL DATA |  |  | BLOOD DATA |  |  |
| --- | --- | --- | --- | --- | --- | --- |
|  | AUC (95%CI) | sensitivity (95%CI) | specificity (95%CI) | AUC (95%CI) | sensitivity (95%CI) | specificity (95%CI) |
| k-Nearest Neighbors | 0.88 (0.75-1.01) | 0.80 (0.55-0.93) | 0.88 (0.66-0.97) | 0.89 (0.78-1) | 0.85 (0.64-0.95) | 0.80 (0.58-0.92) |
| Support Vector Machine (radial) | 0.89 (0.77-1.01) | 0.73 (0.48-0.89) | 0.94 (0.73-0.99) | 0.93 (0.85-1.01) | 0.90 (0.7-0.97) | 0.85 (0.64-0.95) |
| Support Vector Machine (linear) | 0.87 (0.74-1) | 0.87 (0.62-0.96) | 0.82 (0.59-0.94) | 0.93 (0.85-1.01) | 0.90 (0.7-0.97) | 0.85 (0.64-0.95) |
| Partial Least Squares | 0.88 (0.75-1.01) | 0.73 (0.48-0.89) | 0.94 (0.73-0.99) | 0.93 (0.85-1.01) | 0.95 (0.76-0.99) | 0.80 (0.58-0.92) |
| Elastic Net | 0.89 (0.77-1.01) | 0.80 (0.55-0.93) | 0.88 (0.66-0.97) | 0.98 (0.93-1.03) | 0.95 (0.76-0.99) | 0.95 (0.76-0.99) |
| Random Forest | 0.90 (0.78-1.02) | 0.73 (0.48-0.89) | 1.00 (0.82-1) | 0.91 (0.81-1.01) | 0.90 (0.7-0.97) | 0.80 (0.58-0.92) |

All models were trained using the 44 nasal DEGs and 238 blood DEGs on the respective datasets in distinguishing TB from controls. Sensitivity and specificity were estimated at the optimal threshold defined by the Youden index.
DEGs= differentially expressed genes, AUC= area under the curve ; CI= confidence intervals

**Figure 4.**
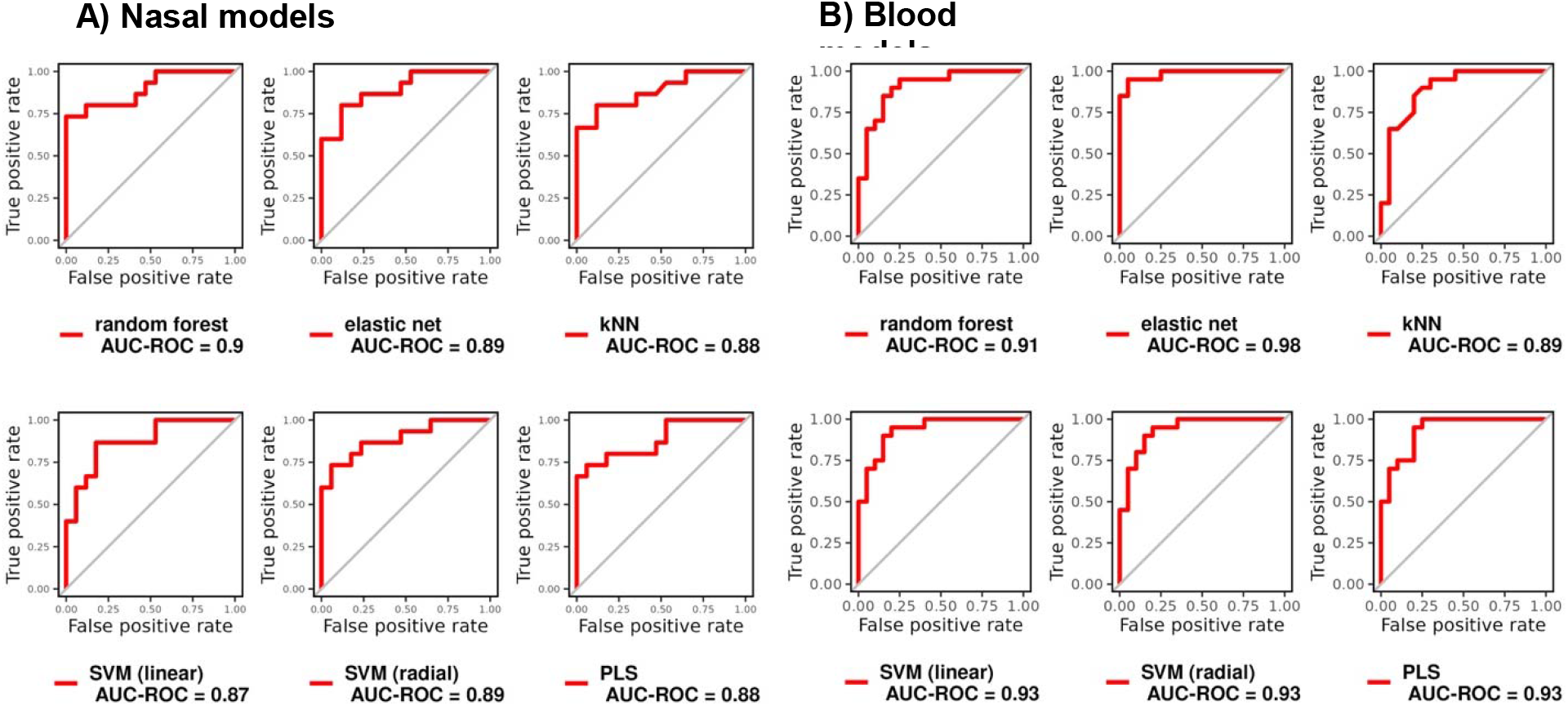
ROC curves and the respective AUC of different machine learning models in distinguishing TB from controls, trained using the (A) 44 nasal DEGs and (B) 238 blood DEGs on the respective datasets, with 10-fold cross-validation repeated 10 times. DEGs= differentially expressed genes, SVM= support vector machine; KNN= k-nearest neighbor, ROC=receiver operating characteristic; AUC= area under the curve, PLS= partial least squares

Amongst the top ten nasal DEGs with the highest variable importance for each model, *SPIB, SHISA2, TESPA1* and *CD1B* were common to all nasal models (**Supplementary Figure 7)**. Models trained using this parsimonious four-gene signature (NASAL_4) had an AUC between 0.87-0.94 for predicting TB status in the nasal data, with 5/6 models meeting the WHO target criteria for a TB triage test (>90% sensitivity and >70% specificity)^4^ **(Supplementary Figure 8 and Supplementary Table 4)**.

### Enrichment pathway analysis

GSEA results are summarized in **Table 2**. The top GSEA enriched nasal pathways were interferon gamma (IFN-γ), neutrophil degranulation and chemokine signaling, all downregulated in adults with TB compared with controls. Enriched downregulated pathways related to cytokine signaling and the inflammatory response, particularly the innate immune system. Upregulated enriched pathways were associated with cilia function, organelle maintenance and cholesterol metabolism. In contrast, in blood, upregulated enriched pathways in TB related to immune activity and hem metabolism, whereas downregulated pathways were associated with transcription and RNA processing. Enriched pathways in interferon and interleukin signaling, neutrophil degranulation, TNF-alpha signaling, complement, inflammatory response, phagocytosis, and leishmania infection were downregulated in nasal samples and upregulated in blood samples amongst adults with TB.

**Table 2:**
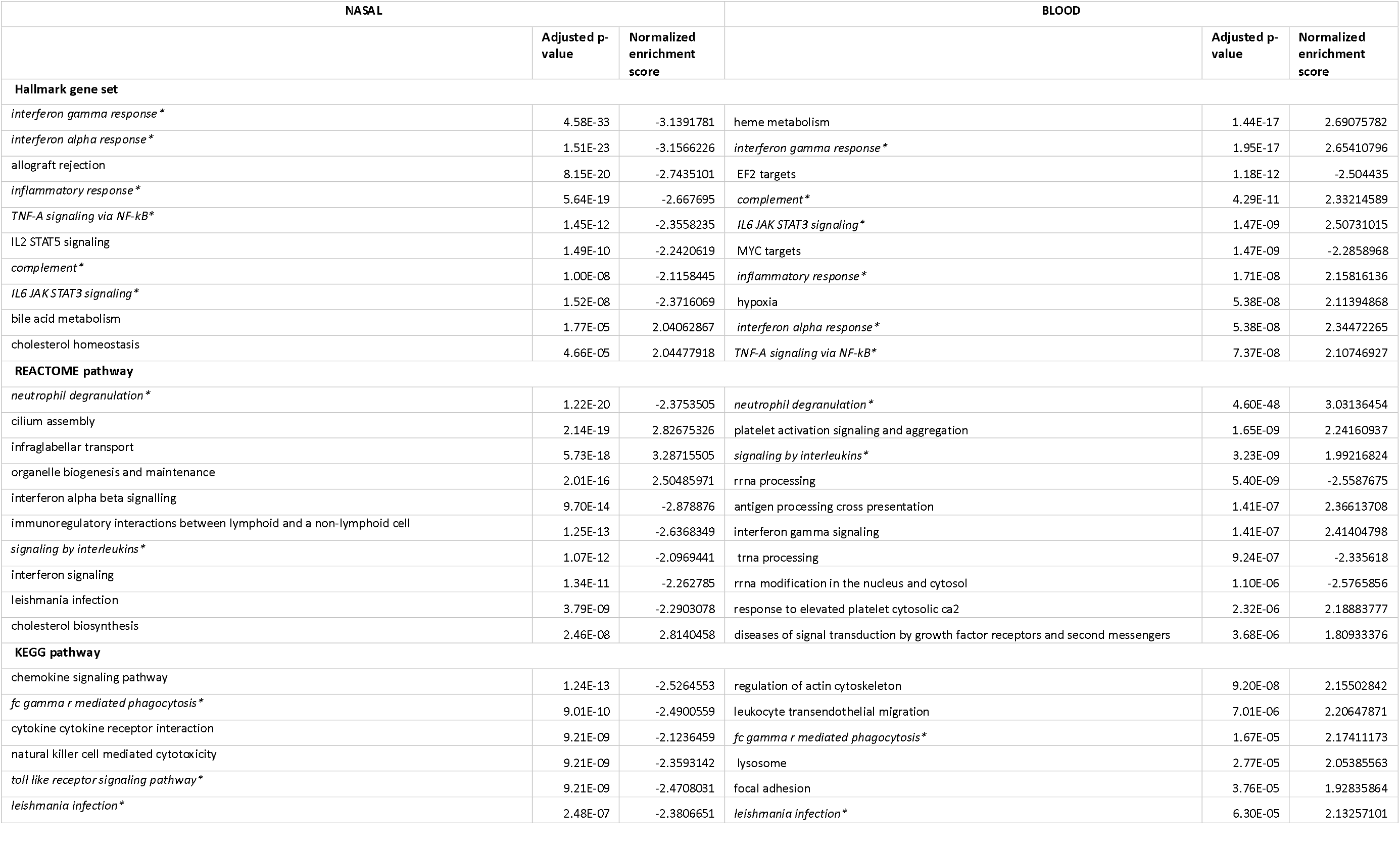
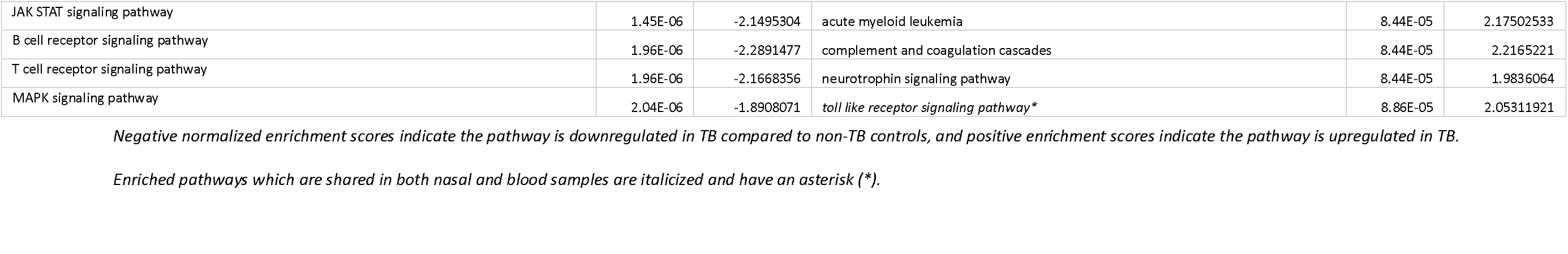
The top ten enriched pathways in the Hallmark gene sets, KEGG and Reactome pathways based on fGSEA in nasal and blood samples.

| NASAL |  |  | BLOOD |  |  |
| --- | --- | --- | --- | --- | --- |
|  | Adjusted p-value | Normalized enrichment score |  | Adjusted p-value | Normalized enrichment score |
| **Hallmark gene set** |  |  |  |  |  |
| *interferon gamma response \** | 4.58E-33 | -3.1391781 | heme metabolism | 1.44E-17 | 2.69075782 |
| *interferon alpha response \** | 1.51E-23 | -3.1566226 | *interferon gamma response \** | 1.95E-17 | 2.65410796 |
| allograft rejection | 8.15E-20 | -2.7435101 | EF2 targets | 1.18E-12 | -2.504435 |
| *inflammatory response \** | 5.64E-19 | -2.667695 | *complement \** | 4.29E-11 | 2.33214589 |
| *TNF-A signaling via NF-kB \** | 1.45E-12 | -2.3558235 | *IL6 JAK STAT3 signaling \** | 1.47E-09 | 2.50731015 |
| IL2 STAT5 signaling | 1.49E-10 | -2.2420619 | MYC targets | 1.47E-09 | -2.2858968 |
| *complement \** | 1.00E-08 | -2.1158445 | *inflammatory response \** | 1.71E-08 | 2.15816136 |
| *IL6 JAK STAT3 signaling \** | 1.52E-08 | -2.3716069 | hypoxia | 5.38E-08 | 2.11394868 |
| bile acid metabolism | 1.77E-05 | 2.04062867 | *interferon alpha response \** | 5.38E-08 | 2.34472265 |
| cholesterol homeostasis | 4.66E-05 | 2.04477918 | *TNF-A signaling via NF-kB \** | 7.37E-08 | 2.10746927 |
| **REACTOME pathway** |  |  |  |  |  |
| *neutrophil degranulation \** | 1.22E-20 | -2.3753505 | *neutrophil degranulation \** | 4.60E-48 | 3.03136454 |
| cilium assembly | 2.14E-19 | 2.82675326 | platelet activation signaling and aggregation | 1.65E-09 | 2.24160937 |
| infraglabellar transport | 5.73E-18 | 3.28715505 | *signaling by interleukins \** | 3.23E-09 | 1.99216824 |
| organelle biogenesis and maintenance | 2.01E-16 | 2.50485971 | rrna processing | 5.40E-09 | -2.5587675 |
| interferon alpha beta signalling | 9.70E-14 | -2.878876 | antigen processing cross presentation | 1.41E-07 | 2.36613708 |
| immunoregulatory interactions between lymphoid and a non-lymphoid cell | 1.25E-13 | -2.6368349 | interferon gamma signaling | 1.41E-07 | 2.41404798 |
| *signaling by interleukins \** | 1.07E-12 | -2.0969441 | trna processing | 9.24E-07 | -2.335618 |
| interferon signaling | 1.34E-11 | -2.262785 | rrna modification in the nucleus and cytosol | 1.10E-06 | -2.5765856 |
| leishmania infection | 3.79E-09 | -2.2903078 | response to elevated platelet cytosolic ca2 | 2.32E-06 | 2.18883777 |
| cholesterol biosynthesis | 2.46E-08 | 2.8140458 | diseases of signal transduction by growth factor receptors and second messengers | 3.68E-06 | 1.80933376 |
| **KEGG pathway** |  |  |  |  |  |
| chemokine signaling pathway | 1.24E-13 | -2.5264553 | regulation of actin cytoskeleton | 9.20E-08 | 2.15502842 |
| *fc gamma r mediated phagocytosis \** | 9.01E-10 | -2.4900559 | leukocyte transendothelial migration | 7.01E-06 | 2.20647871 |
| cytokine cytokine receptor interaction | 9.21E-09 | -2.1236459 | *fc gamma r mediated phagocytosis \** | 1.67E-05 | 2.17411173 |
| natural killer cell mediated cytotoxicity | 9.21E-09 | -2.3593142 | lysosome | 2.77E-05 | 2.05385563 |
| *toll like receptor signaling pathway \** | 9.21E-09 | -2.4708031 | focal adhesion | 3.76E-05 | 1.92835864 |
| *leishmania infection \** | 2.48E-07 | -2.3806651 | *leishmania infection \** | 6.30E-05 | 2.13257101 |
| JAK STAT signaling pathway | 1.45E-06 | -2.1495304 | acute myeloid leukemia | 8.44E-05 | 2.17502533 |
| B cell receptor signaling pathway | 1.96E-06 | -2.2891477 | complement and coagulation cascades | 8.44E-05 | 2.2165221 |
| T cell receptor signaling pathway | 1.96E-06 | -2.1668356 | neurotrophin signaling pathway | 8.44E-05 | 1.9836064 |
| MAPK signaling pathway | 2.04E-06 | -1.8908071 | *toll like receptor signaling pathway\** | 8.86E-05 | 2.05311921 |

**575** *Negative normalized enrichment scores indicate the pathway is downregulated in TB compared to non-TB controls, and positive enrichment scores indicate the pathway is upregulated in TB.*
**576** *Enriched pathways which are shared in both nasal and blood samples are italicized and have an asterisk (\*).*

Using ORA, no enriched nasal pathways were identified with an adjusted p-value of <0.05. With an adjusted p-value of <0.1, upregulated nasal DEGs in TB pertained mainly to metabolic biological processes and cell division and differentiation whereas inflammatory and immune pathways were enriched in the downregulated genes **(Supplementary Figure 9A, 9C)**. In blood, upregulated DEGs were enriched in the inflammatory and immune response, whereas biological processes in the downregulated DEGs included synaptic transmission, odontogenesis and regulation of cell migration (adjusted p-value <0.05) **(Supplementary Figure 9B, 9D)**.

Canonical pathways in IPA also demonstrated suppressed immune activity in nasal samples from adults with TB, including reduced interferon signaling, phagosome formation, and S100 family and G-protein-coupled receptor signaling. In contrast, blood showed activation of innate immune pathways, such as neutrophil degranulation, IFN-γ, toll-like receptor signaling, inflammasomes, pyroptosis and iNOS signaling **(Supplementary Figure 10A, 10B)**. This opposing pattern of inflammation and interferon response was supported by upstream regulator analysis which predicted inhibition of type I and type III interferon expression (IFNA2, IFNL1), interferon regulatory factors (IRF1), JAK-STAT components and pathogen-recognition receptors (TLR9) in the nose and activation of inflammatory regulators (IFN-γ, CSF2, IL5, TNF, IL15, TLR2, SELPG) in blood **(Supplementary Figure 10C, 10D)**. Overall, GSEA, ORA and IPA suggested there was reduced nasal mucosal inflammation and increased systemic inflammation in TB disease.

***Table 2 should appear here***. Table 2 is larger than one A4 page in length and as per BMC Infectious Diseases Submission Guidelines is found at the end of ocument text file.

### Immune cell composition

Enrichment analysis suggested that the immune response, particularly the innate immune system, was downregulated in the nasal samples and upregulated in blood in adults with TB. Heterogeneity in immune cell composition was explored using CIBERSORTX and LM22. There were no statistically significant differences in the proportions of immune cell subtypes in nasal samples based on TB status. However, for blood, the proportion of neutrophils was higher in TB (adjusted p-value = 0.0376), whereas natural killer cells were lower in TB (adjusted p-value = 0.0376). Other types of immune cells in blood showed no significant differences **(Supplementary Figure 11)**.

### TB Signature Profiler

In the RNA sequencing dataset, only 6/59 published TB diagnostic blood signatures distinguished TB from controls with an AUC ≥0.7 and p-value <0.05 in the nasal dataset (Maertzdorf_100, Lee_5, Sloot_2, Natarajan_7, Jacobensen_3 and Gjoen_10^46^) **(Figure 5A, 5C)**. In contrast, most published blood signatures distinguished TB from controls in our blood dataset (44/59 and 41/59 had an AUC ≥0.7 and p-value <0.05 using ssGSEA and GSVA respectively) **(Figure 5B, 5D)**. Only one signature which performed well in the nasal dataset (Jacobsen_3) performed well in the blood dataset. Full AUC results are in **Supplementary Table 5 and 6**.

**Figure 5.**
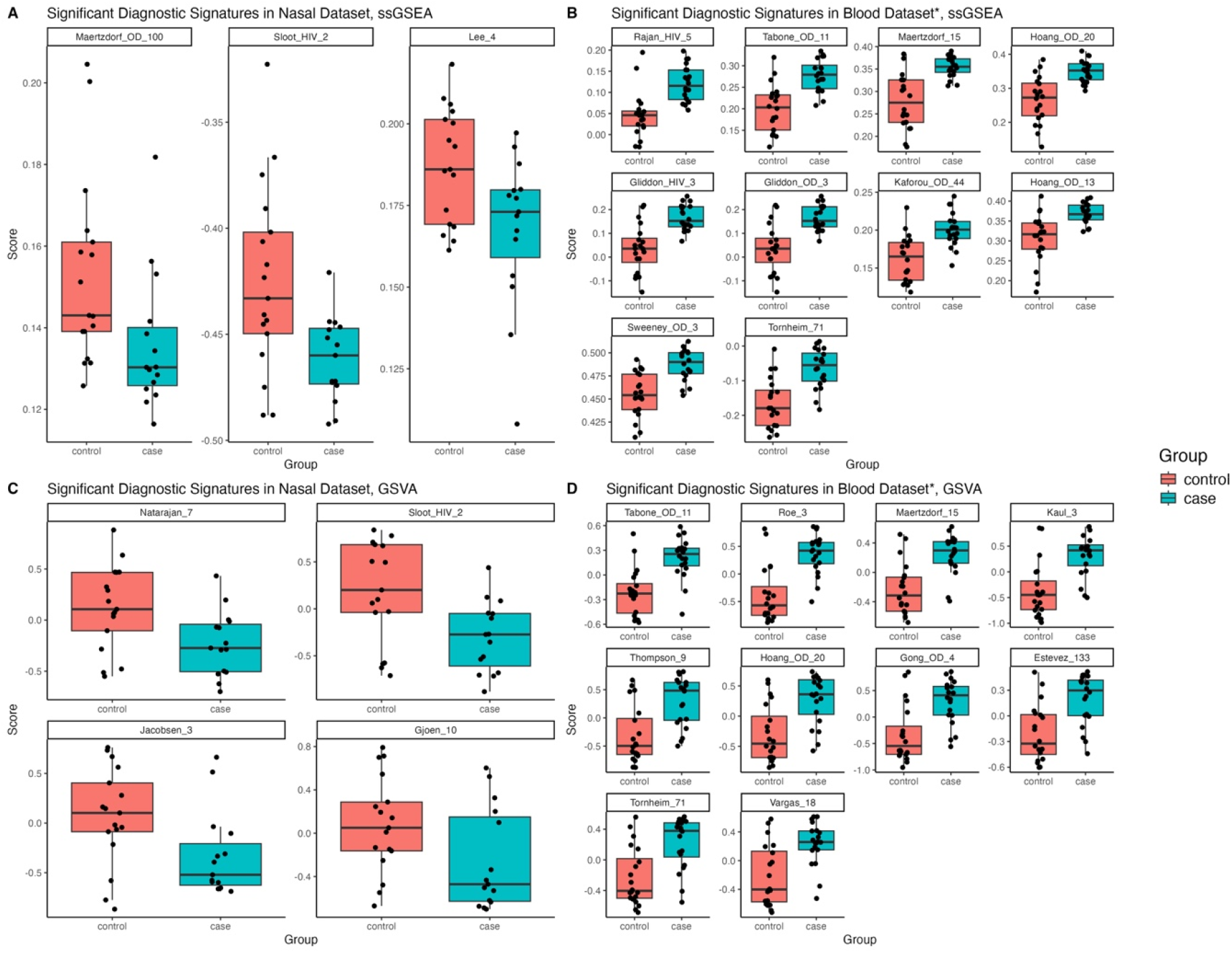
Boxplots of published blood diagnostic signatures that reached statistical significance (p value <0.05) and had a bootstrapped AUC of >0.7 in distinguishing TB from controls using ssGSEA in the A) nasal and B) blood datasets and using GSVA methods in the C) nasal and D) blood datasets. *For blood, only the top ten (based on AUC) published blood diagnostic signatures are shown. GSVA= gene set variation analysis; ssGSEA= single-sample gene set enrichment analysis.

## Discussion

In a cohort of treatment-naive PLHIV with presumptive TB in Uganda, we identified 44 DEGs in nasal cells between adults with microbiologically confirmed TB disease compared with individuals without TB. These 44 DEGs yielded a promising AUC of > 0.87 in predicting TB across six models in cross-validation within our dataset. Amongst those with TB disease, we also found a blunted inflammatory nasal response, with downregulation of several innate immune pathways. Compartmental differences were seen, with contrasting patterns of inflammation in TB between the nose and blood and little overlap in nasal and blood DEGs. Our proof-of-concept study highlights that there are differences in nasal gene expression associated with TB disease that are distinct from blood.

There are no published nasal transcriptomic studies in TB; however, half of the top ten nasal DEGs have been associated with TB in blood. *SPIB*, which ranked amongst the most important predictive genes across all models, is important for plasmacytoid dendritic cell function^47^ and memory B cell maintenance^48^. *SPIB* downregulation was noted in active TB patients in multiple studies^49-51^. *CLC*, which is linked to neutrophil recruitment^52^, was part of a four-marker blood signature for differentiating adults with TB and non-TB febrile infection^53^. *ITLN*, which has a role in pathogen recognition, was downregulated in newly diagnosed adults with TB, increasing in expression with treatment^49^. Changes in expression of T cell markers *PTGDR2* and *CD69* have also been associated with early signs of pulmonary TB^54^ and TB infection, respectively^55^. Top DEGs not previously linked to TB include *CEACAM21*, which facilitates opsonin-independent recognition and clearance of specific gram-negative bacterial pathogens^56^. *CST1, CLC, FETUB* and *VSMT1* were upregulated in asthma and allergic diseases in nasal studies^57-59^, which could reflect overrepresentation of allergic diseases in our non-TB control group.

GSEA, ORA and IPA suggested that TB was associated with suppressed nasal innate immune responses. IFN-γ activity was the top enriched downregulated pathway from the Hallmark gene-sets using fGSEA. IFN-γ is an important cytokine for immunity against mycobacteria. It facilitates the host defense to *Mtb* through stimulating macrophage activity and producing reactive nitrogen species^60^, with ineffective IFN-γ activity associated with disseminated mycobacterial infection^61^. Neutrophils were the top downregulated REACTOME enriched pathway. Whilst excessive neutrophil activation can cause tissue damage, they have a protective role in early stages^62^ and adult contacts with higher blood neutrophil counts were shown to be less likely to develop TB infection^63^. Chemokine signaling was the most enriched downregulated KEGG pathway. Single nucleotide polymorphisms in chemokines involved in cell migration in TB, were shown to increase susceptibility to active TB^64,65^. The nose is the first line of defense for airborne pathogens and early control of respiratory infection is dependent on robust nasal mucosal immune responses^66^. Dampening of IFN-γ activity, neutrophils, and chemokines in addition to other enriched nasal pathways like complement, phagocytosis, and toll-like receptor signaling, may underlie failure of the early protective nasal response to *Mtb* in TB disease.

The nasal host response in TB differed from blood. Few published TB blood signatures could classify TB in our nasal sequencing dataset. Moreover, downregulated nasal innate immune responses in TB were accompanied by upregulation of inflammatory pathways in blood. Unlike blood, nasal samples have more heterogeneity in cell composition, consisting of both immune and epithelial components^14^. We could not show that nasal immune pathways were driven by immune cell composition. However, neutrophils in the blood were higher in PLHIV with TB versus those without TB, consistent with our observation of increased neutrophil enrichment in blood.

Our finding of upregulated innate immune pathways in TB blood signatures confirms previously published studies in PLHIV^10,67,68^. Similar to the downregulation of immune nasal pathways in TB in our study, Mick and colleagues found that toll-like receptor signaling, inflammasomes, neutrophil degranulation and chemokine signaling were attenuated in the nasopharyngeal response in SARS-CoV-2 patients compared to other viral infections^16^. Hurst and colleagues also noted that systemic symptoms in SARS-CoV-2-infected individuals were correlated with suppressed nasal innate and adaptive immune pathways and upregulation of these same pathways in matched blood samples^69^. In contrast, two studies comparing nasal and blood samples in children with viral infections, identified upregulation in immune activity and strong innate responses in both compartments^14,15^. Although our cross-sectional study cannot infer causality, we can hypothesize that reduced mucosal immune responses and inadequate control of *Mtb* in the nose could promote progression to lung disease and consequent systemic inflammation. Longitudinal studies using paired nasal and blood samples are needed to understand the temporal relationship between the nasal and blood immune response and TB disease.

A streamlined nasal gene signature based on variable importance (NASAL_4) met the WHO criteria for a TB triage test (>90% sensitivity and 70% specificity) in cross-validation. Three genes within NASAL_4 relate to immune activity: *SPIB* is linked to interferon production and proinflammatory cytokines^47^, *CD1b* is associated with antigen presentation^70^ and *TESPA* relates to T cell maturation^71^. Whilst the AUC for nasal models based on all DEGs was slightly lower than blood, most of the top ten nasal DEGs were not ISGs, unlike blood. This is promising given that the performance of many ISG-dominant RNA blood signatures in differentiating TB from viral infections or in PLHIV has been inadequate^8,10^. Moreover, adults with TB missed by blood models were classified correctly by nasal models and vice versa, suggesting potential value of combined nasal-blood diagnostic biomarkers.

All adults in our study had HIV. Studies of the nasal transcriptome in HIV are limited, however HIV causes mucosal immune dysfunction^72^, which will shape nasal transcriptional changes, independently of TB. Based on flow cytometry and single cell sequencing, Phiri and colleagues found increased nasal neutrophilic abundance and inflammation in PLHIV compared with non-infected individuals and that nasal neutrophils had transcriptional signatures of stress and senescence and nasal T cells had signatures of exhaustion and apoptosis^73^. Differing numbers of nasal CD3 and CD4 cells based on HIV status have also been noted^74^. Further studies looking at the nasal TB response among individuals without HIV would help explore the extent to which nasal gene profiles in our study are driven by HIV.

Our work has limitations. The reliance on sputum Ultra as the TB reference standard in PLHIV means paucibacillary TB disease could be missed in the control group. We could not follow up controls to identify later development of TB disease and were unable to evaluate the role of nasal signatures in predicting TB progression. Secondly, our ART-naive cohort was at high risk of opportunistic infections and nasopharyngeal bacterial colonization. However, we were unable to ascertain and adjust for these as well as any alternative diagnoses in non-TB controls that would affect gene expression. Case numbers were too small to study the impact of CD4 count on nasal signatures, and other covariates that impact gene expression, like malnutrition and TB infection via TST or IGRA status, were unavailable. Finally, nasal DEG-based classifiers were trained and evaluated on the same dataset since the sample size was too small to allocate a separate test set, and there are no other published nasal transcriptomic datasets for TB. Although we employed k-fold cross-validation to reduce overfitting, the need for external validation of nasal signatures on independent datasets remains.

Despite these limitations, these are the first data interrogating the nasal transcriptome in TB disease. Whilst developing a nasal diagnostic test will require a more accessible technology than sequencing, detecting gene expression differences is the first step. Our focus on ART-naïve PLHIV who have an elevated risk of TB addresses a major gap in HIV-TB studies and by only studying symptomatic adults, we remove the bias of using healthy controls. Although outside the scope of this work, sequencing nasal samples enable the concurrent study of the respiratory microbiome and opportunity to develop integrated host-microbiome diagnostic biomarkers. Although we focused on adults diagnosed with Ultra on expectorated sputum as our microbiological reference standard, a nasal biomarker could still have additional value for TB diagnosis given the ease of sampling compared with sputum. Future work to confirm the utility of nasal signatures in TB requires discovery and external validation of nasal biomarkers across larger, independent, geographically distinct cohorts, that include adults with and without HIV across the spectrum of TB disease, as well as other difficult to diagnose groups, such as young children or those without microbiologically confirmed TB.

## Conclusion

In conclusion, our exploratory study using minimally invasive nasal samples suggests that host nasal transcriptomics is feasible and informative for the diagnosis of TB, whereby elucidating gene expression profiles at the initial pathogen encounter reveals novel insights into host-TB interaction. We found distinct nasal gene expression patterns associated with TB disease in PLHIV that were not captured in blood, with DEGs able to distinguish TB from non-TB controls. Our findings support further study of the nose for non-sputum-based TB diagnostic biomarkers, which is an urgent research priority to reduce global TB-associated mortality.

<u>Additional tables greater than one A4 in length</u>

## Data Availability

Computational code and data are available at: https://github.com/nisreenkhambati/uganda_nasal_cells. 
Raw expression data for nasal and blood samples  are publicly available on the SRA database (PRJNA1379661) on https://www.ncbi.nlm.nih.gov/bioproject/PRJNA1379661

https://www.ncbi.nlm.nih.gov/bioproject/PRJNA1379661

https://github.com/nisreenkhambati/uganda_nasal_cells

## List of abbreviations

ART: antiretroviral therapy
AUC: area under the curve
DEG: differentially expressed genes
GSEA: gene set enrichment analysis
GSVA: gene set variation analysis
IPA: ingenuity pathway analysis
KNN: k nearest neighbors
ORA: over representation analysis
PCA: principal component analysis
PLHIV: people living with HIV
PLS: partial least squares
RF: random forest
ROC: receiver operating characteristic curve
SVM: support vector machine
ssGSEA: single sample gene set enrichment analysis
TB: tuberculosis
WHO: world health organization

## Declarations

### Ethics approval and consent to participate

This study used human data. Informed consent was obtained from all participants. The study protocol and informed consent forms was approved, with ethics approval given by institutional review boards of all partnering institutions (Boston University Medical Campus, Joint Clinical Research Centre and Uganda National Council for Science and Technology). This study did not use any animal data.

### Consent for publication

Not applicable

### Availability of data and materials

The datasets analyzed during the current study are available in the SRA repository (PRJNA1379661) on https://www.ncbi.nlm.nih.gov/bioproject/PRJNA1379661 Computational code and datasets are also available at: https://github.com/nisreenkhambati/uganda_nasal_cells.

### Author contributions

Conceptualization: JE, LN, WEJ, SG

Methodology: NK, KLT, LN, WEJ

Resources: LN, WEJ, PS

Project administration: LN, WEJ

Investigation: EN, LN, KLT, NK

Software: NK

Data curation: EN, NK

Formal analysis: NK (except IPA done by AVV)

Validation: KLT, SL, JKA, WEJ

Writing–original draft: NK

Writing –review of final manuscript: all authors

Visualisation-NK

Supervision: RS, AJP, EMB, DOC, WEJ

Funding acquisition: JE, LN, WEJ, SG

## Acknowledgements

We would like to thank all the participants in Uganda who were part of this study, as well as all local clinical, lab and data teams involved.

