## Supplementary Figures and Tables for "Nasal Gene Expression in ART-Naive Adults with HIV and Pulmonary Tuberculosis in Uganda"

|  | **Total cohort**  **(HIV+)**  **n=40** | **Adults with TB (HIV+/TB+)**  **n=20** | **Controls**  **(HIV+/TB-)**  **n=20** | **p-value** |
| --- | --- | --- | --- | --- |
| Sex |  |  |  |  |
| Female, n (%) | 22 (55) | 9 (45) | 13 (65) | 0.34 |
| Male, n (%) | 18 (45) | 11 (55) | 7 (35) |  |
| Median age, (IQR), years | 34 (27-38) | 33 (27-38) | 35 (27-40) | 0.56 |
| Median CD4 count, (IQR), cells/mm^3^ | 182 (92-346) | 207 (118-331) | 162 (42 -349) | 0.61 |
| CD4 count less than 100 |  |  |  |  |
| Yes, n (%) | 10 (25) | 4 (20) | 6 (30) | 0.72 |
| No, n (%) | 30 (75) | 16 (80) | 14 (70) |  |
| HIV stage |  |  |  |  |
| Stage 1, n (%) | 3 (7.5) | 0 | 3 (15) | 0.11 |
| Stage 2, n (%) | 15 (37.5) | 10 (50) | 5 (25) |  |
| Stage 3, n (%) | 22 (55) | 10 (50) | 12 (60) |  |
| Ultra semiquantitative result if TB diagnosed |  |  |  |  |
| Very low, n (%) | 1 (5)* | 1 (5) | - | NA |
| Low n (%) | 7 (35)* | 7 (35) | - |  |
| Medium, n (%) | 3 (15)* | 3 (15) | - |  |
| High, n (%) | 9 (45)* | 9 (45) | - |  |

*Supplementary Table 1: Baseline characteristics among TB and controls, with comparison of variables highlighted by p values. Continuous variables were compared using the Student’s t-test or Mann-Whitney U test , whereby the normality of the data distribution was first checked using the Shapiro-Wilk Normality Test. Categorical variables were compared using the Pearson's chi-square or Fisher’s exact test (if expected cell counts <5) . Differences in groups with p-values <0.05 were deemed statistically significant.*

*•percentage of Ultra semiquantitative result based on adults with TB (n=20) rather than total cohort*

| **Quality indicator** | **Nasal** | **Blood** |
| --- | --- | --- |
| Proportion of reads uniquely mapped to human genome, median (IQR), % | 67 (44-74) | 57 (50-64) |
| Reads per sample, median (IQR), millions | 16.5 (11.1- 18.7) | 7.9 (6.6-9.2) |

*Supplementary Table 2: Quality metrics of nasal and blood sequencing alignment using the Spliced Transcripts Alignment to a Reference (STAR) tool*

*IQR: interquartile range*

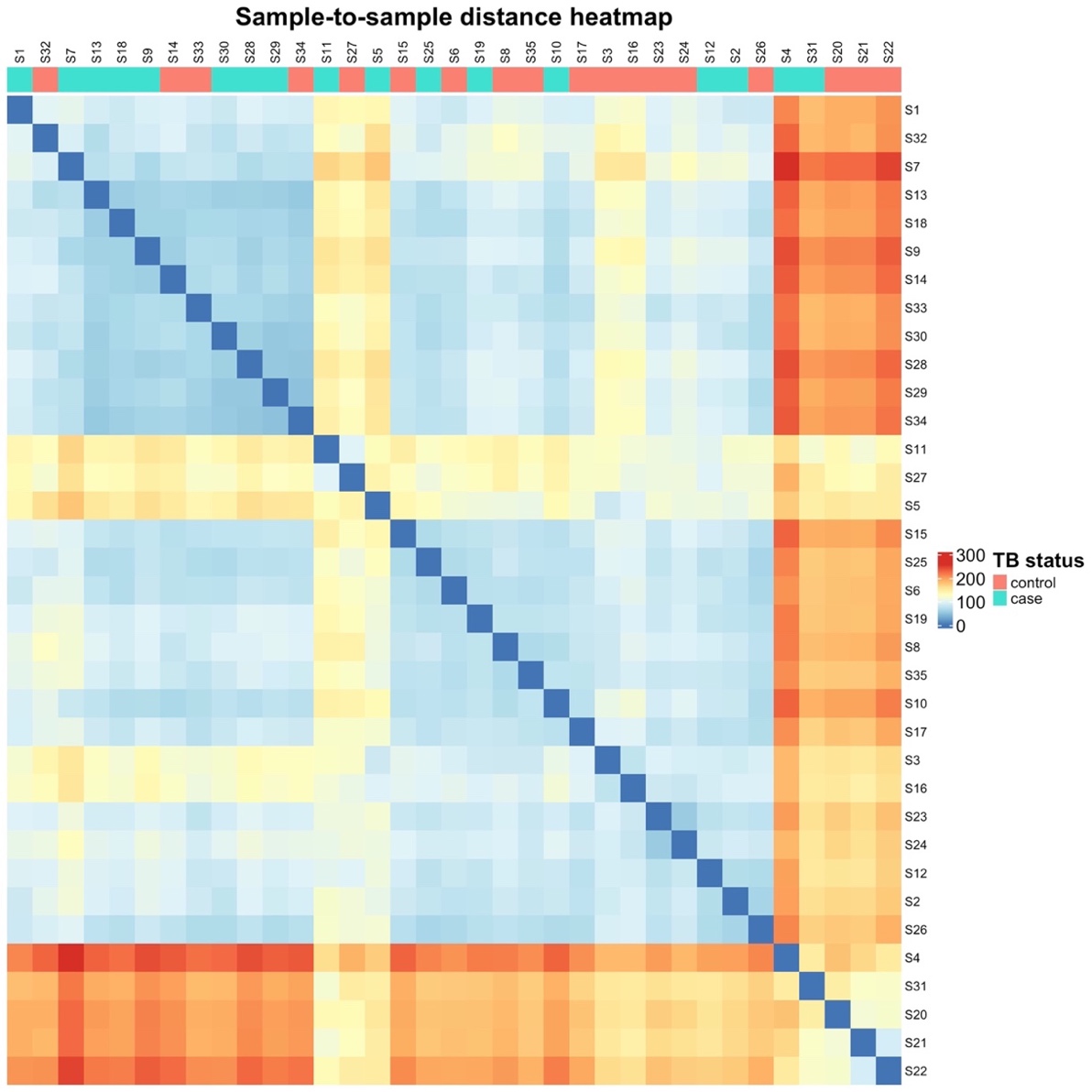

*Supplementary Figure 1: Heatmap based on sample-sample Euclidean distance for all pairwise combinations of nasal samples. Five samples (S4, S22, S31, S20, S21) demonstrated high average sample-to-sample distances with most other samples, irrespective of sample TB status. Three of these samples (S4, S20, S31) had less than 5 million reads and were excluded from further downstream analysis.*

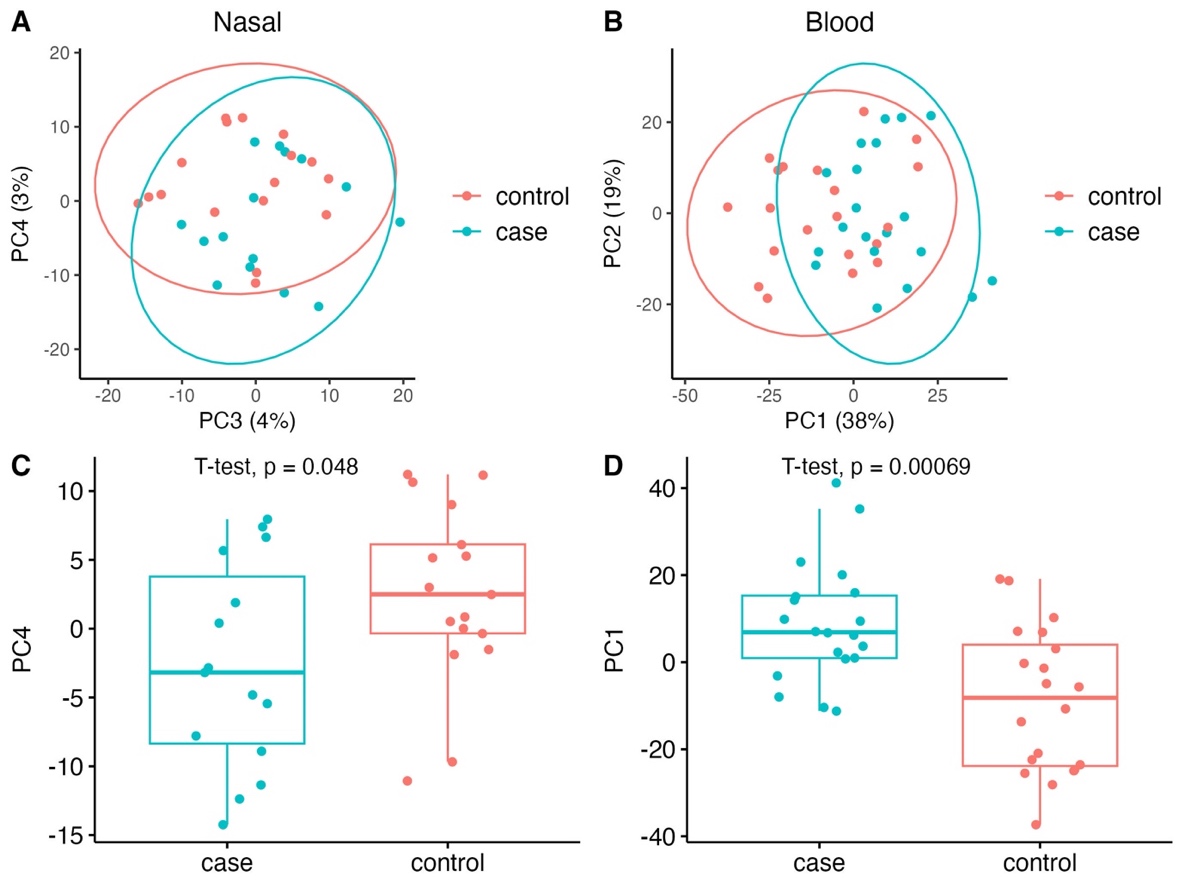

*Supplementary Figure 2: PCA and boxplots of nasal and blood gene expression, based on TB status.*

*A: PCA of nasal sample gene expression for PC3 and PC4, with 95% confidence ellipses.*

*B: PCA of blood gene expression for PC1 and PC2, with 95% confidence ellipses.*

*C:  Boxplots demonstrating differences in PC4 between TB and controls for nasal samples, using a two-sided student’s T-test.*

*D:  Boxplots demonstrating differences in PC1 between TB cases and controls for blood samples, using a two-sided student’s T-test.*

*PCA = principal component analysis; PC1= principal component 1; PC2= principal component 2; PC3= principal component 3; PC4= principal component 4.*

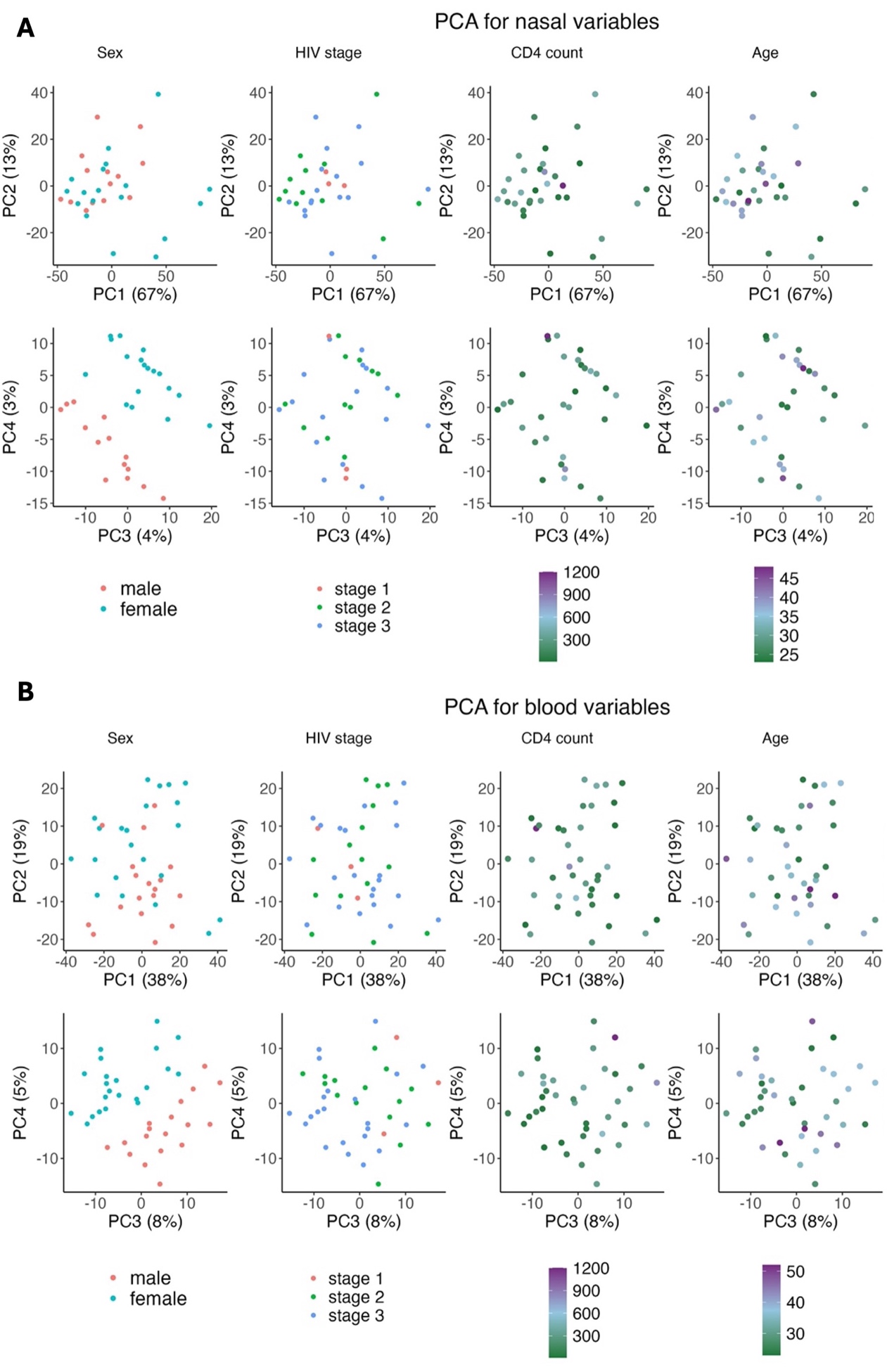

*Supplementary Figure 3: PCA of sex, HIV-related variables and age for A) nasal and B) blood samples. Males clustered separately from females on PC3 and PC4 in nasal and blood samples*

*PCA = principal component analysis; PC1= principal component 1; PC2= principal component 2; PC3= principal component 3; PC4= principal component 4.*

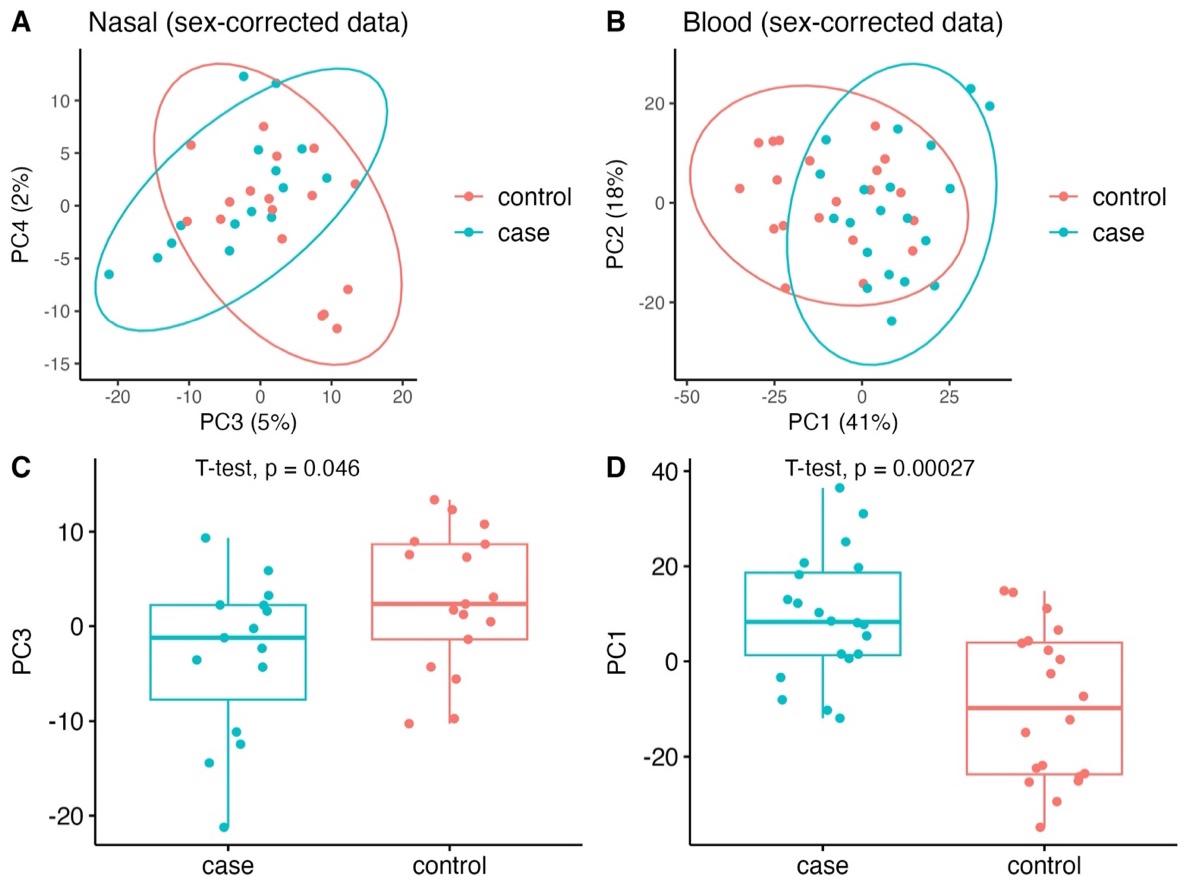

*Supplementary Figure 4: PCA and boxplots of nasal and blood gene expression based on TB status, following correction for sex.*

*A: PCA of nasal sample gene expression for PC3 and PC4, with 95% confidence ellipses.*

*B: PCA of blood gene expression for PC1 and PC, with 95% confidence ellipses.*

*C:  Boxplots demonstrating statistically significant differences in PC3 between TB cases and controls for nasal samples, using a two-sided Student’s T-test.*

*D:  Boxplots demonstrating statistically significant differences in PC1 between TB cases and controls for blood samples, using a two-sided Student’s T-test.*

*PCA = principal component analysis; PC1= principal component 1; PC2= principal component 2; PC3= principal component 3; PC4= principal component 4.*

| **Nasal DEG (n=44)** | **Blood DEG (n=238)** | | | |
| --- | --- | --- | --- | --- |
| CST1 | GBP6 | FKBP5 | RNF208 | CYP27A1 |
| CLC | APOL4 | LTF | KCNJ15 | ABCA13 |
| LGALS12 | NRN1 | CEACAM1 | TMEM119 | MINDY4B |
| ITLN1 | SLC6A19 | PLIN4 | ITGA2B | TCL1A |
| SPIB | VPS9D1 | MAPK14 | ADM | TCN1 |
| FETUB | COL4A2 | HP | HMGA2 | ADRA2A |
| CEACAM21 | SEPTIN4 | RETN | MCEMP1 | PADI4 |
| CD69 | GBP5 | ZNF438 | C1QTNF7 | CD300LD |
| PTGDR2 | SORT1 | CCL2 | VPREB3 | AJAP1 |
| VSTM1 | IL18RAP | KREMEN1 | OLFM4 | PRSS33 |
| CRISP2 | ADCY3 | CTNNAL1 | HRK | GLDN |
| CLEC12A | ATP1B2 | TNNT3 | S100A12 | SIGLEC11 |
| CEBPE | LHFPL2 | RALGAPA2 | SCN9A | PADI2 |
| CCR3 | FCGR1A | CDCP1 | OLAH | TIMM10 |
| SHISA2 | MED12L | UGT2B28 | S100A9 | CFAP97D1 |
| TPSAB1 | IL18R1 | BATF2 | KCNH7 | CMTM5 |
| TESPA1 | GPR84 | EFCAB2 | CACNA2D3 | CCN2 |
| CISH | MAOB | CERS1 | CEL | C1QA |
| CD1B | RGS16 | ZDHHC19 | CD200 | CCIN |
| ADAM19 | PHTF1 | RAB20 | ADGRG3 | MGAM |
| SIGLEC6 | BEND7 | TP63 | CLU | G0S2 |
| PRSS33 | PSTPIP2 | ALOX15 | PAX9 | SHOC1 |
| SLC9A3 | MAP3K20 | BCL6 | PTGDR2 | BICDL2 |
| SORD | PFKFB2 | CMBL | SLC36A3 | ERICH3 |
| OTOF | CD274 | RCVRN | SEMA6B | BMP3 |
| CLEC12B | AIM2 | ACE2 | MYO10 | OR2T8 |
| GAS1 | SCARF1 | ARG1 | ITGB3 | CRISP3 |
| BTNL8 | MYOF | NECTIN2 | SPNS3 | SPTA1 |
| FHL3 | CD177 | CEACAM8 | CR1 |  |
| POSTN | ANKRD22 | SOCS3 | VIT |  |
| IFIT2 | INSL3 | LIPM | FGFR2 |  |
| ZBTB16 | SHROOM4 | SIPA1L2 | GALNT14 |  |
| FABP6 | BTBD19 | METTL7B | AQP1 |  |
| RSAD2 | GRB10 | PFKFB3 | KLHDC8A |  |
| CCR7 | NAIP | KCNH8 | CRISPLD2 |  |
| CASS4 | TIFA | PCP4L1 | KCNH2 |  |
| DYDC1 | NUPR1 | SRGAP3 | TMEM92 |  |
| GAPT | MMRN1 | LIMK2 | GATA6 |  |
| IL31RA | CASP5 | ALDH1A2 | GNG10 |  |
| CNR2 | KAZN | PLXNB3 | ACSL1 |  |
| CDH26 | PDCD1LG2 | RMI2 | ANKRD9 |  |
| HS3ST3A1 | MMP8 | MMP9 | HSD3B7 |  |
| COL28A1 | SELP | TMEM132D | LONRF3 |  |
| HRH4 | ADAMTS2 | SLC29A1 | CACNG8 |  |
|  | PCOLCE2 | SMARCD3 | TRIM9 |  |
|  | OPLAH | BCAM | SNX7 |  |
|  | ELAPOR1 | IGF2BP2 | CKAP4 |  |
|  | UGCG | BPI | ANXA3 |  |
|  | ITGA7 | DYSF | AXL |  |
|  | CACNA1E | SCN1B | CLDN18 |  |
|  | BMX | ERG | C1QB |  |
|  | IRAK3 | SPTB | LILRA4 |  |
|  | LOXL1 | KCNS2 | PGLYRP1 |  |
|  | GADD45G | RD3L | TAS2R40 |  |
|  | TPST1 | CNGB1 | CAMP |  |
|  | ASPH | CREB5 | CCN3 |  |
|  | IL1R2 | LRRC17 | SIGLEC8 |  |
|  | DSC2 | COL23A1 | NOS1AP |  |
|  | P2RY14 | MARCO | RAP1GAP |  |
|  | DNAH17 | PLIN5 | CYP2J2 |  |
|  | GK | CC2D2A | VNN1 |  |
|  | RAPGEF5 | PROS1 | C4BPA |  |
|  | FLT3 | EGF | PRKAG3 |  |
|  | GNLY | CALHM6 | NRXN1 |  |
|  | SPATC1 | DYRK3 | ZFHX4 |  |
|  | CACNB4 | CCNA1 | CLC |  |
|  | HTRA3 | LRG1 | IL1R1 |  |
|  | SERPING1 | CLRN1 | TDRD9 |  |
|  | GBP1 | S100A8 | DUSP13 |  |
|  | CLEC5A | HPN | SLC26A8 |  |

*Supplementary Table 3: Table of 44 nasal DEGS and 238 blood DEGs, based on differential gene expression analysis between PLHIV and TB disease and PLHIV without TB disease.*

*DEG = differentially expressed genes; PHIV: people living with HIV*

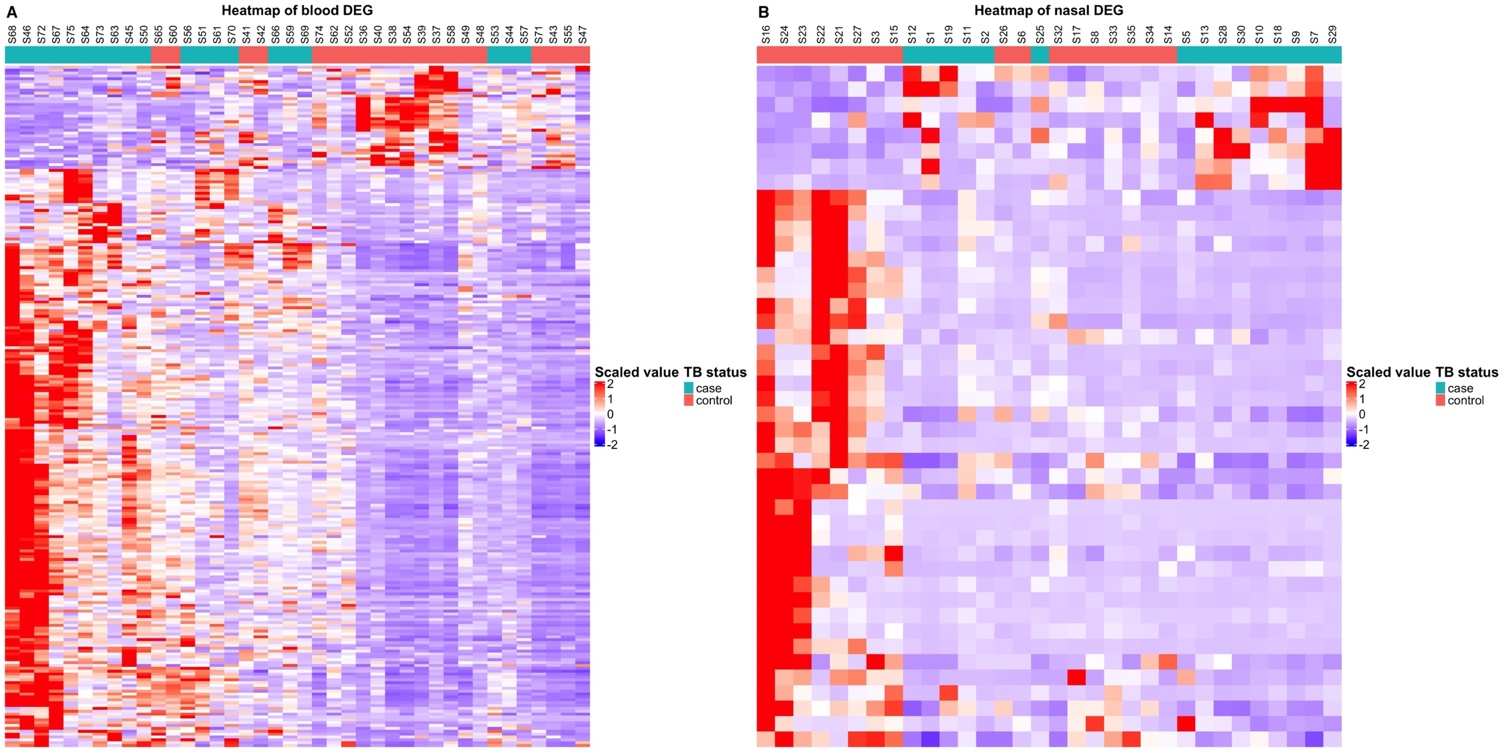

*Supplementary Figure 5: Heatmap based on (A) 238 blood DEGs and (B) 44 nasal DEGs, showing clustering of TB cases and controls in the respective datasets. The columns represent the samples, and the rows represent the DEGs. Sample clustering used Euclidean distance as a measure of dissimilarity and complete linkage for between-cluster separation.*

*DEG = differentially expressed genes*

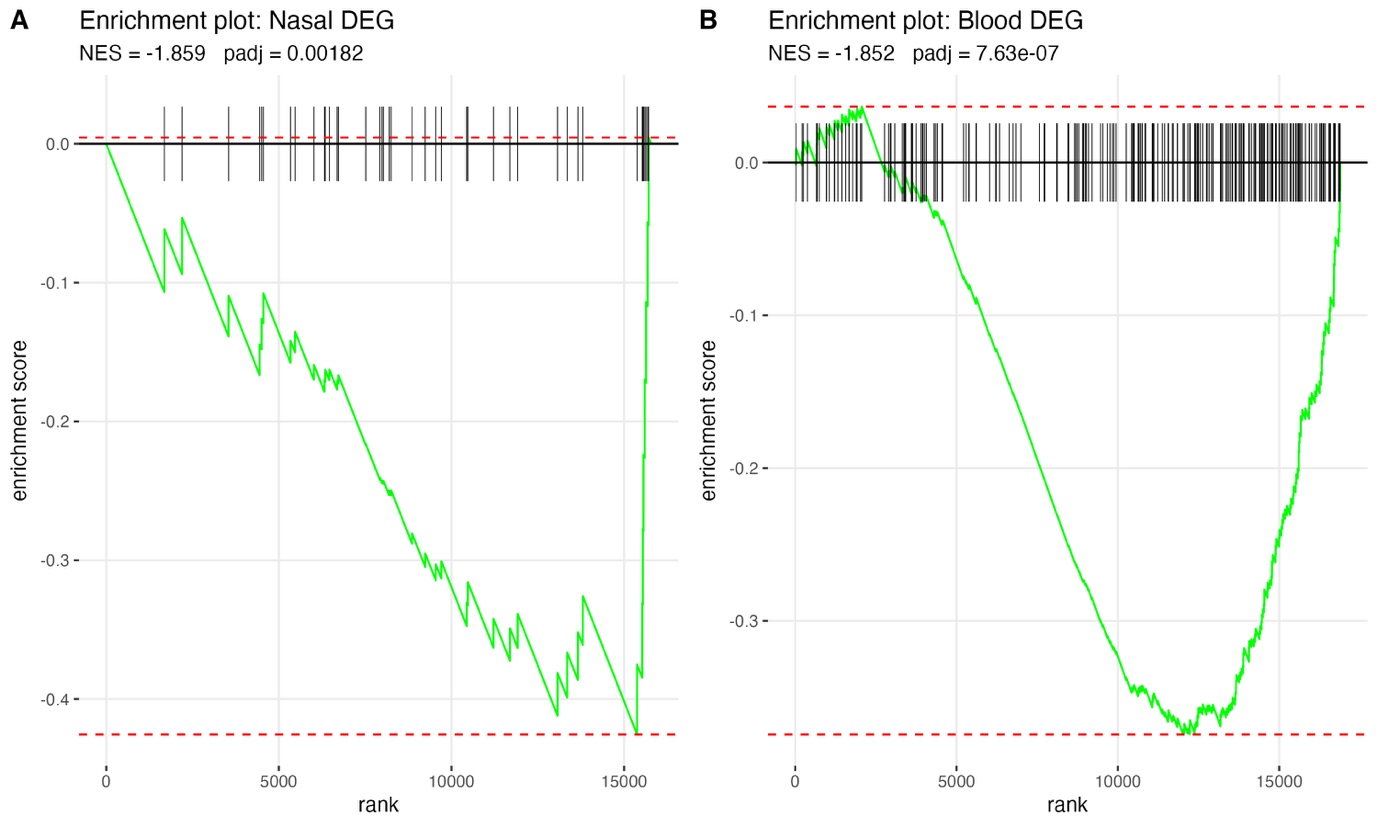

*Supplementary Figure 6: Enrichment plots from FGSEA for A) the nasal DEG gene set in the ranked blood gene list and B) the blood DEG gene set in the ranked nasal gene list. Vertical black lines represent positions of the DEGs within the ranking and the green line represents the running enrichment score. Both tissue DEGs were significantly negatively enriched in the other tissue ranked lists, suggesting nasal immune responses in TB differ from blood, with transcriptional changes occurring in opposite directions.*

*DEG = differentially expressed genes; NES: normalized enrichment score; padj: adjusted p values*

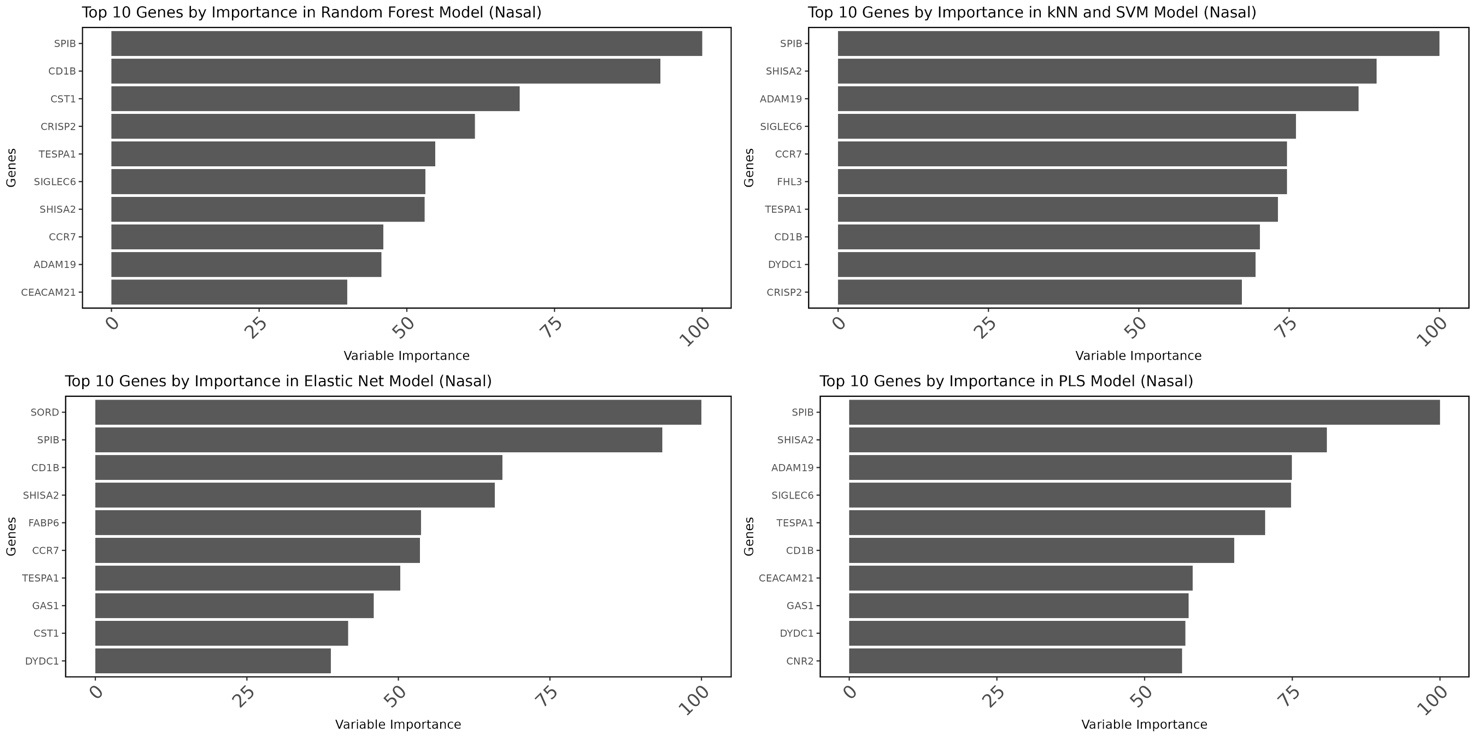
*Supplementary Figure 7: The top ten genes ranked by variable importance in the nasal models. All variable importance scores were scaled to have a maximum value of 100. For SVM and KNN classification models, no built-in importance score is implemented, therefore the area under the ROC curve for each variable was used as the measure of importance.*

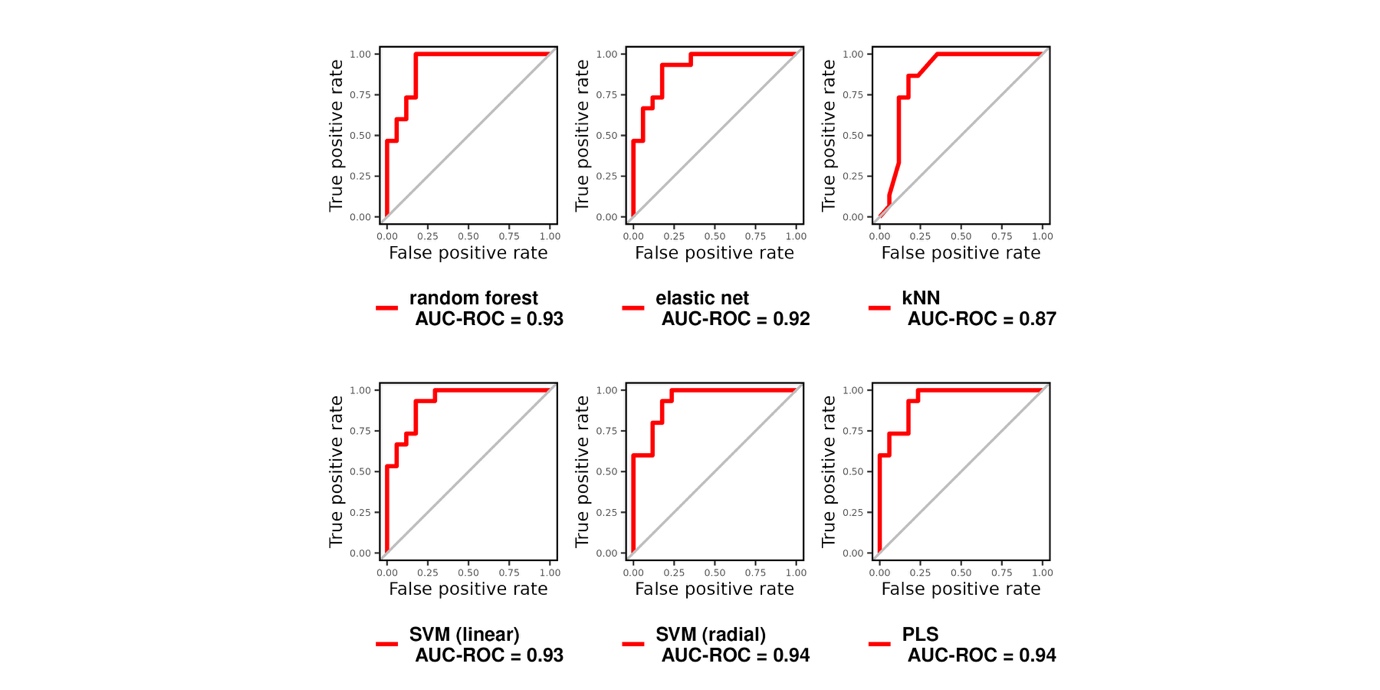

**ROC curves of NASAL_4**

*Supplementary Figure 8: ROC curves and the respective AUC of machine learning models in distinguishing TB from controls, trained using the NASAL_4 signature (SPIB, SHISA2, TESPA1, CD1B) with 10-fold cross-validation repeated 10 times.*

*DEGs= differentially expressed genes, SVM= support vector machine; KNN= k-nearest neighbor, ROC=receiver operating characteristic; AUC= area under the curve, PLS= partial least squares*

|  | **NASAL_4 signature** | | |
| --- | --- | --- | --- |
|  | **AUC**  **(95%CI)** | **sensitivity**  **(95%CI)** | **specificity**  **(95%CI)** |
| **k-Nearest Neighbors** | 0.87 (0.74-1) | 0.86 (0.62-0.96) | 0.82 (0.59-0.94) |
| **Support Vector Machine (radial)** | 0.94 (0.85-1.03) | 1.00 (0.8-1) | 0.77 (0.53-0.9) |
| **Support Vector Machine (linear)** | 0.93 (0.83-1.03) | 0.93 (0.7-0.99) | 0.82 (0.59-0.94) |
| **Partial Least Squares** | 0.94 (0.85-1.03) | 1.00 (0.8-1) | 0.77 (0.53-0.9) |
| **Elastic Net** | 0.92 (0.82-1.02) | 0.93 (0.7-0.99) | 0.82 (0.59-0.94) |
| **Random Forest** | 0.93 (0.83-1.03) | 1.00 (0.8-1) | 0.82 (0.59-0.94) |

*Supplementary Table 4: Diagnostic performance with 95% CI of each model, trained using NASAL_4 (SPIB, SHISA2, TESPA1, CD1B) with 10-fold cross-validation repeated 10 times, in distinguishing TB from controls. Sensitivity and specificity were estimated at the optimal threshold defined by the Youden index.*

*DEGs= differentially expressed genes, AUC= area under the curve ; CI= confidence intervals*

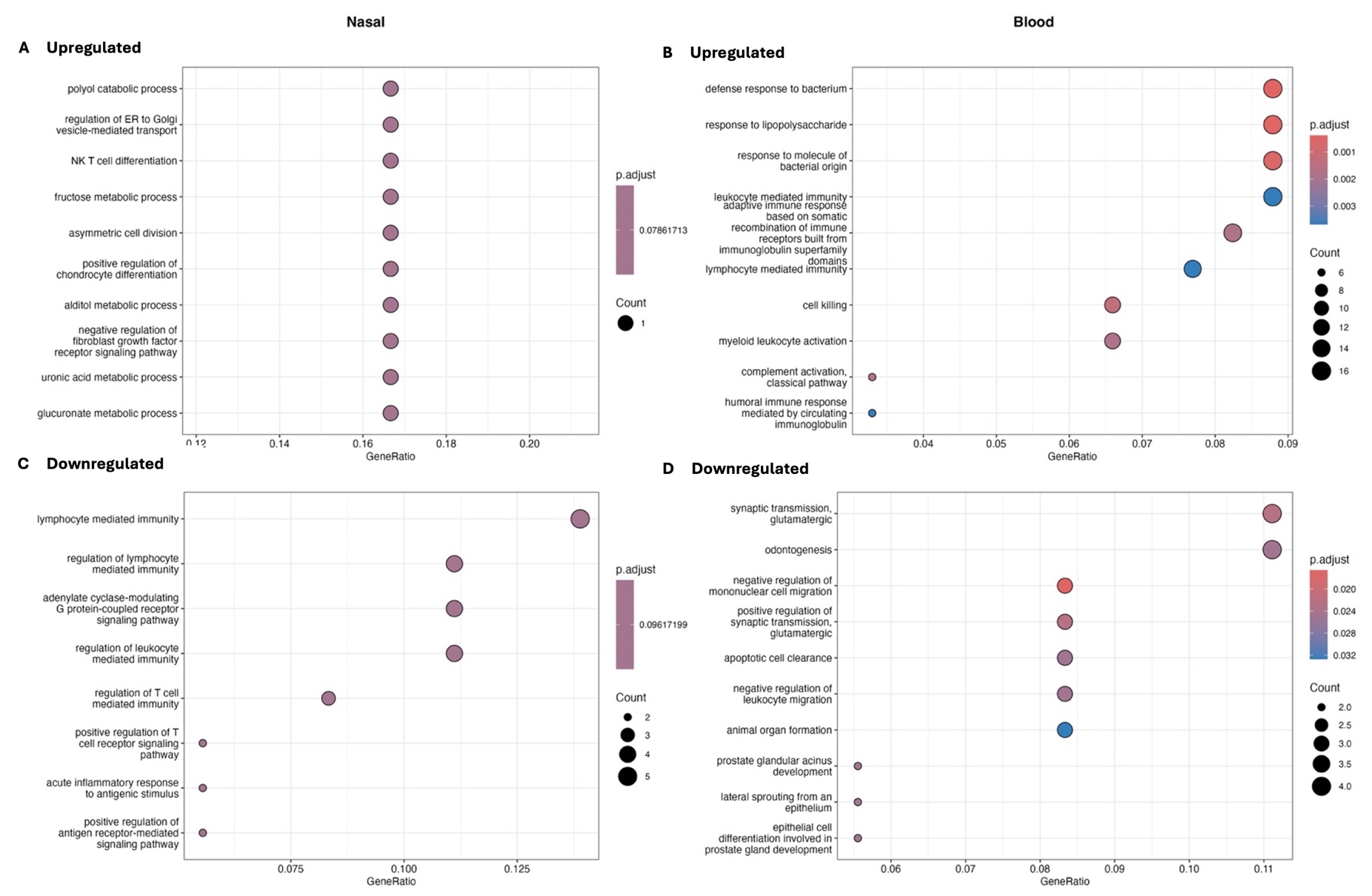

*Supplementary Figure 9: Dot plots of ORA showing the top significant GO biological processes enriched in A) upregulated nasal DEGs, B) upregulated blood DEGs, C) downregulated nasal DEGs, and D) downregulated blood DEGs. Gene ratio is the fraction of DEGs belonging to an ontology over the total number of DEGs. Circle size shows the number of DEGs per biological process. DEGs= differentially expressed genes; ORA= overrepresentation analysis.*

*.*

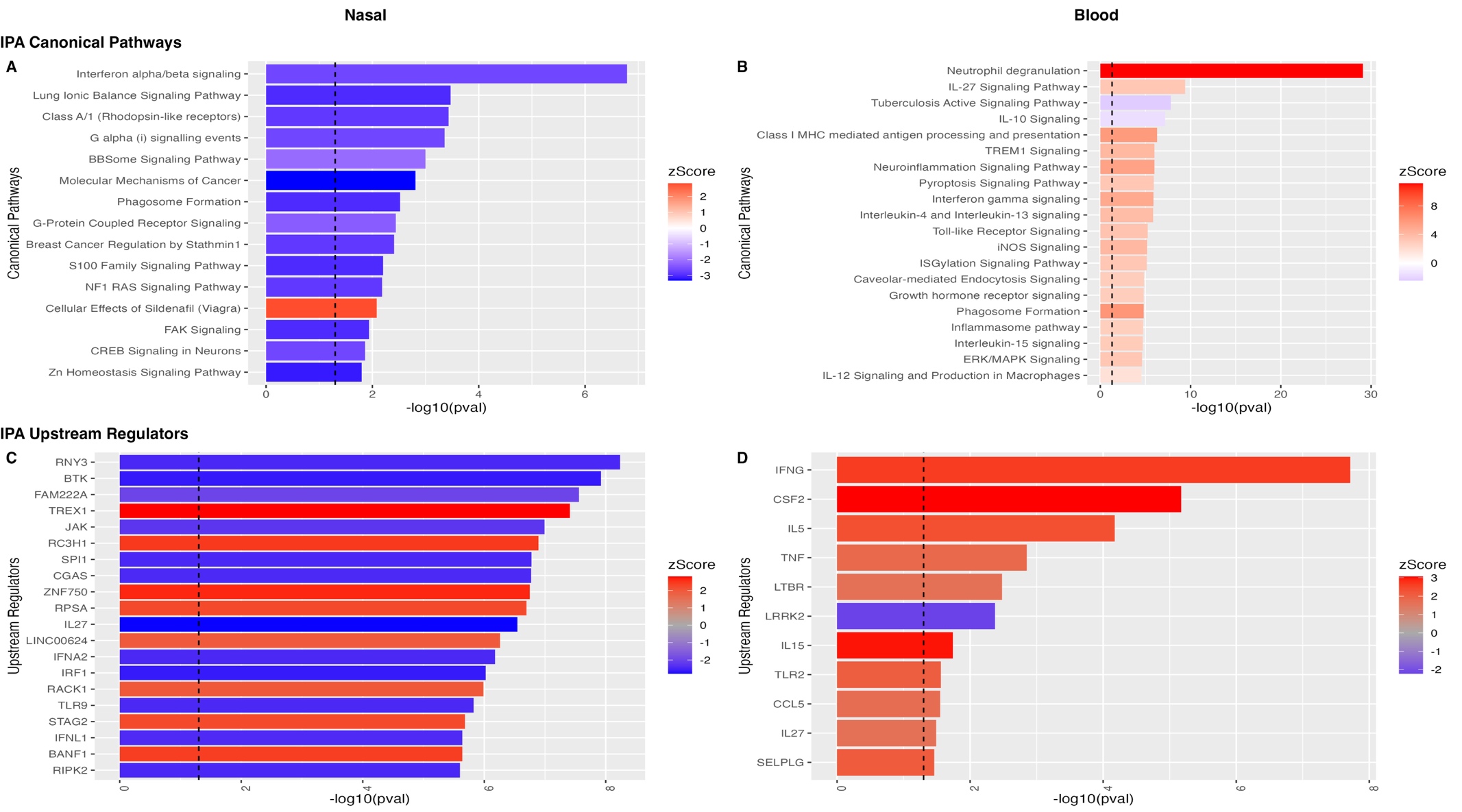

*Supplementary Figure 10: Top Ingenuity Pathway Analysis canonical pathways in nasal (A) and blood (B) samples and upstream regulators for nasal (C) and blood (D) samples in TB, ranked by significance(-log10pvalue). Bar color represents the IPA activation z-score: pathways and regulators in blue are inhibited in adults with TB, whereas those in red are activated in TB. The dashed vertical line represents the IPA significance threshold (p-value= 0.05). Only pathways with p-values <0.05 and IPA z-score≥ ±1.5 are shown, filtered to a maximum of top 20 pathways.*

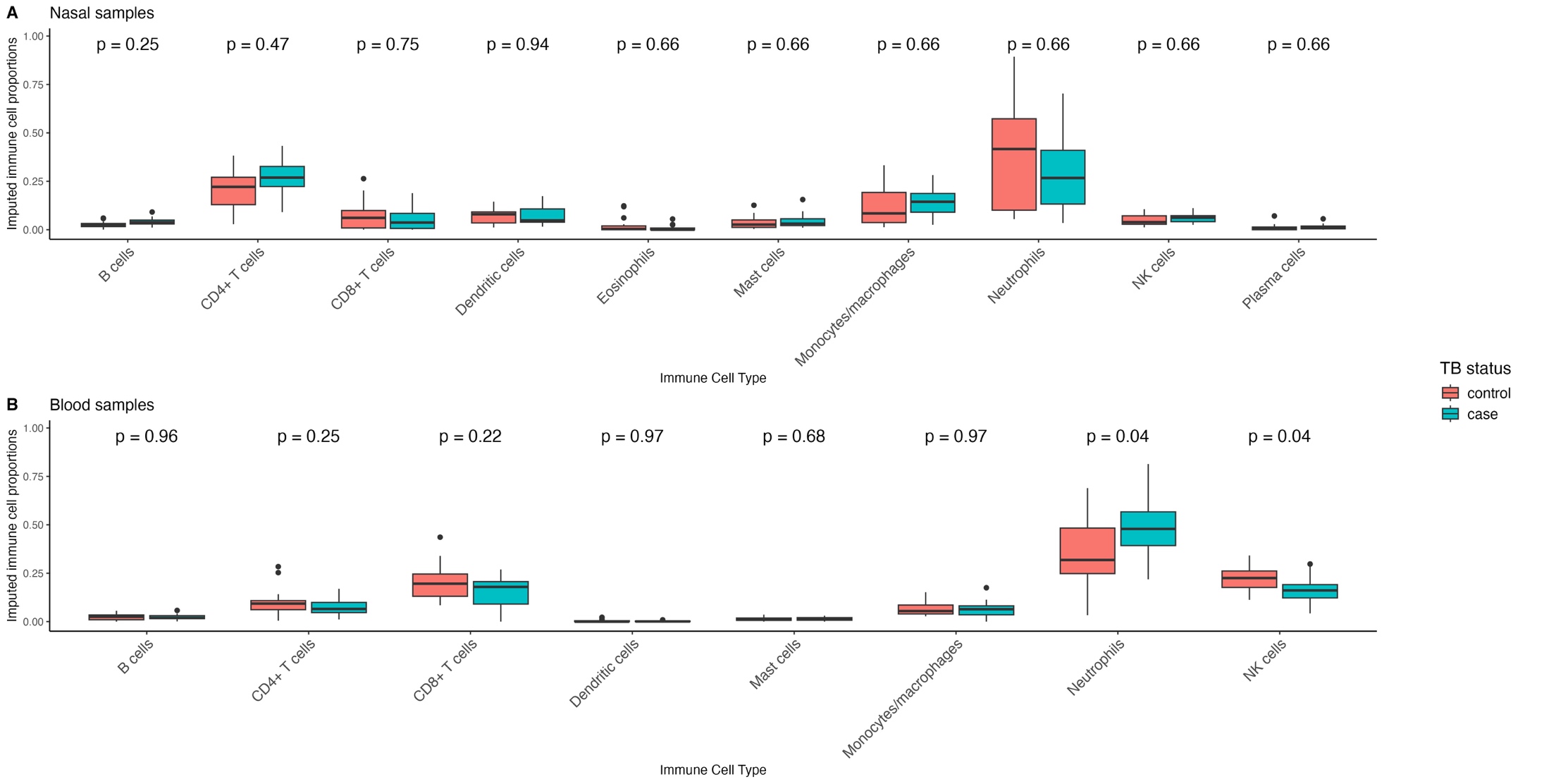

*Supplementary Figure 11:  Boxplots of proportions of immune cell populations in A) nasal and B) blood samples, imputed using CIBERSORTX. All 32 nasal and 40 blood samples had a p-value < 0.05 for the deconvolution and were included in the analysis. Proportions of immune cells were compared by TB status using either Student’s t-test or Mann-Whitney U test , with all analyses adjusted for multiple testing. Adjusted p values are shown above the boxplots. Only immune cell populations present in at least 50% of samples are shown.*

|  | **Nasal dataset** | | | | | | | |
| --- | --- | --- | --- | --- | --- | --- | --- | --- |
|  | **ssGSEA** | | | | **GSVA** | | | |
| **Signature** | **P value** | **Lower AUC 95%CI** | **AUC** | **Upper AUC 95% CI** | **P value** | **Lower AUC 95% CI** | **AUC** | **Upper AUC 95% CI** |
| **Anderson_42** | 0.9755 | 0.5 | 0.5059 | 0.7436 | 0.4201 | 0.5061 | 0.5608 | 0.7653 |
| **Anderson_OD_51** | 0.4626 | 0.5042 | 0.5725 | 0.7471 | 0.6266 | 0.5063 | 0.5529 | 0.7899 |
| **Berry_393** | 0.0986 | 0.5324 | 0.6588 | 0.8314 | 0.173 | 0.509 | 0.6314 | 0.7623 |
| **Berry_OD_86** | 0.0908 | 0.502 | 0.6471 | 0.7835 | 0.1609 | 0.5051 | 0.651 | 0.8179 |
| **Blankley_380** | 0.1319 | 0.5098 | 0.6549 | 0.8286 | 0.1774 | 0.5206 | 0.651 | 0.8758 |
| **Blankley_5** | 0.0831 | 0.5118 | 0.6588 | 0.8493 | 0.293 | 0.5059 | 0.6039 | 0.7838 |
| **Bloom_OD_144** | 0.2084 | 0.506 | 0.6235 | 0.822 | 0.4432 | 0.5078 | 0.6039 | 0.8078 |
| **Chen_5** | 0.118 | 0.506 | 0.6157 | 0.8101 | 0.5625 | 0.5061 | 0.5569 | 0.7462 |
| **Chen_HIV_4** | 0.4332 | 0.5022 | 0.5333 | 0.7261 | 0.6581 | 0.504 | 0.5412 | 0.724 |
| **Dawany_HIV_251** | 0.0499 | 0.5093 | 0.6706 | 0.832 | 0.6277 | 0.5079 | 0.5608 | 0.7626 |
| **Duffy_23** | 0.122 | 0.5098 | 0.6275 | 0.8058 | 0.3421 | 0.5106 | 0.5961 | 0.7814 |
| **Esmail_203** | 0.0984 | 0.5088 | 0.6745 | 0.8532 | 0.2129 | 0.5058 | 0.6392 | 0.8549 |
| **Esmail_82** | 0.2078 | 0.5098 | 0.6353 | 0.8447 | 0.4183 | 0.502 | 0.6078 | 0.7742 |
| **Esmail_OD_893** | 0.1188 | 0.5182 | 0.6588 | 0.8311 | 0.1718 | 0.511 | 0.6471 | 0.8389 |
| **Estevez_133** | 0.1452 | 0.5103 | 0.6431 | 0.8358 | 0.2289 | 0.5137 | 0.6471 | 0.8305 |
| **Estevez_259** | 0.2329 | 0.5068 | 0.5961 | 0.829 | 0.441 | 0.5054 | 0.5804 | 0.7804 |
| **Francisco_OD_2** | 0.089 | 0.5275 | 0.6824 | 0.8541 | 0.1833 | 0.5081 | 0.6275 | 0.8084 |
| **Gjoen_10** | 0.0593 | 0.5283 | 0.702 | 0.8865 | 0.0494 | 0.5274 | 0.7059 | 0.8429 |
| **Gjoen_7** | 0.1002 | 0.5142 | 0.6941 | 0.867 | 0.155 | 0.5025 | 0.6588 | 0.8364 |
| **Gliddon_2_OD_4** | 0.9644 | 0.506 | 0.5255 | 0.7436 | 0.4769 | 0.5029 | 0.5725 | 0.74 |
| **Gliddon_HIV_3** | 0.6276 | 0.5023 | 0.5294 | 0.7495 | 0.5697 | 0.5059 | 0.5333 | 0.7392 |
| **Gliddon_OD_3** | 0.6276 | 0.503 | 0.5294 | 0.7258 | 0.5697 | 0.5039 | 0.5333 | 0.7112 |
| **Gliddon_OD_4** | 0.9644 | 0.5059 | 0.5255 | 0.7247 | 0.4769 | 0.5081 | 0.5725 | 0.7758 |
| **Gong_OD_4** | 0.0618 | 0.5374 | 0.6706 | 0.8529 | 0.1364 | 0.5079 | 0.6784 | 0.8199 |
| **Hoang_OD_13** | 0.043 | 0.5164 | 0.6784 | 0.8431 | 0.1877 | 0.5117 | 0.6314 | 0.8172 |
| **Hoang_OD_20** | 0.0871 | 0.5169 | 0.6471 | 0.8306 | 0.2979 | 0.5088 | 0.6157 | 0.8148 |
| **Huang_OD_13** | 0.2722 | 0.518 | 0.6431 | 0.8009 | 0.2933 | 0.5079 | 0.6196 | 0.8127 |
| **Jacobsen_3** | 0.1016 | 0.5342 | 0.702 | 0.8702 | 0.0196 | 0.5714 | 0.7451 | 0.8998 |
| **Jenum_8** | 0.1678 | 0.5127 | 0.6588 | 0.8203 | 0.0554 | 0.5157 | 0.6784 | 0.8333 |
| **Kaforou_27** | 0.2876 | 0.5029 | 0.5686 | 0.7706 | 0.1133 | 0.5193 | 0.6588 | 0.8288 |
| **Kaforou_OD_44** | 0.4099 | 0.5022 | 0.502 | 0.7544 | 0.3489 | 0.5074 | 0.5882 | 0.8018 |
| **Kaforou_OD_53** | 0.1373 | 0.5228 | 0.6667 | 0.8277 | 0.0524 | 0.5218 | 0.698 | 0.8729 |
| **Kaul_3** | 0.0503 | 0.5228 | 0.6902 | 0.8601 | 0.1075 | 0.5031 | 0.6588 | 0.8128 |
| **Kulkarni_HIV_2** | 0.3762 | 0.5097 | 0.5961 | 0.7825 | 0.47 | 0.5069 | 0.5882 | 0.793 |
| **Kwan_186** | 0.1873 | 0.5078 | 0.6235 | 0.7904 | 0.2643 | 0.5097 | 0.6039 | 0.8279 |
| **LauxdaCosta_OD_3** | 0.0963 | 0.5263 | 0.698 | 0.8743 | 0.1168 | 0.5147 | 0.6745 | 0.8654 |
| **Lee_4** | 0.0137 | 0.5468 | 0.7333 | 0.892 | 0.1712 | 0.5317 | 0.6784 | 0.8401 |
| **Leong_24** | 0.1746 | 0.5069 | 0.6314 | 0.793 | 0.7703 | 0.5071 | 0.5137 | 0.7579 |
| **Maertzdorf_15** | 0.0542 | 0.5374 | 0.7098 | 0.871 | 0.1844 | 0.5079 | 0.6392 | 0.7999 |
| **Maertzdorf_4** | 0.0961 | 0.5105 | 0.651 | 0.8508 | 0.1289 | 0.5182 | 0.6902 | 0.8439 |
| **Maertzdorf_OD_100** | 0.0236 | 0.5876 | 0.7843 | 0.9219 | 0.1108 | 0.5342 | 0.6706 | 0.8347 |
| **Natarajan_7** | 0.0972 | 0.5223 | 0.698 | 0.8707 | 0.0105 | 0.6219 | 0.7608 | 0.9054 |
| **Qian_OD_17** | 0.1265 | 0.5195 | 0.6275 | 0.7711 | 0.5239 | 0.5055 | 0.549 | 0.7368 |
| **Rajan_HIV_5** | 0.2757 | 0.5089 | 0.6431 | 0.8149 | 0.349 | 0.5197 | 0.6275 | 0.798 |
| **Roe_3** | 0.0832 | 0.511 | 0.6863 | 0.8599 | 0.1096 | 0.5115 | 0.6667 | 0.887 |
| **Roe_OD_4** | 0.0739 | 0.5166 | 0.6471 | 0.8095 | 0.0432 | 0.5424 | 0.698 | 0.8567 |
| **Sambarey_HIV_10** | 0.2261 | 0.505 | 0.5961 | 0.7874 | 0.2083 | 0.5083 | 0.6275 | 0.8142 |
| **Singhania_OD_20** | 0.3897 | 0.5048 | 0.6078 | 0.7892 | 0.6419 | 0.5039 | 0.5333 | 0.73 |
| **Sivakumaran_11** | 0.069 | 0.5521 | 0.702 | 0.884 | 0.1425 | 0.5156 | 0.6275 | 0.8219 |
| **Sloot_HIV_2** | 0.0056 | 0.614 | 0.7765 | 0.9087 | 0.0061 | 0.5906 | 0.7725 | 0.9216 |
| **Sweeney_OD_3** | 0.0732 | 0.5147 | 0.6824 | 0.8681 | 0.2341 | 0.5042 | 0.6353 | 0.8232 |
| **Tabone_OD_11** | 0.0747 | 0.5309 | 0.6863 | 0.8388 | 0.1391 | 0.5039 | 0.6706 | 0.879 |
| **Thompson_9** | 0.2314 | 0.5176 | 0.6588 | 0.8679 | 0.4597 | 0.5137 | 0.6078 | 0.7993 |
| **Tornheim_71** | 0.7257 | 0.502 | 0.5451 | 0.7532 | 0.6414 | 0.502 | 0.5647 | 0.7764 |
| **Vargas_18** | 0.054 | 0.517 | 0.6784 | 0.8744 | 0.1478 | 0.5079 | 0.6549 | 0.8209 |
| **Vargas_42** | 0.0677 | 0.5276 | 0.6627 | 0.8269 | 0.1389 | 0.5219 | 0.6431 | 0.7819 |
| **Verhagen_10** | 0.3428 | 0.5059 | 0.6039 | 0.8222 | 0.22 | 0.5059 | 0.6196 | 0.8045 |
| **Walter_51** | 0.2646 | 0.5192 | 0.6118 | 0.7997 | 0.7219 | 0.507 | 0.5686 | 0.7938 |
| **Zhao_NANO_6** | 0.1209 | 0.51 | 0.6314 | 0.8253 | 0.2996 | 0.5136 | 0.6275 | 0.8557 |

*Supplementary Table 5: Results from the TBSignature Profiler evaluating the diagnostic performance of 59 published blood signatures in classifying TB status in the nasal sequencing dataset. Two scoring methods were used: GSVA and ssGSEA. p-values were calculated using a 2-sample t-test. AUC values are provided with confidence intervals derived from bootstrapping. GSVA= gene set variation analysis; ssGSEA= single-sample GSEA, CI= confidence intervals.*

|  | **Blood dataset** | | | | | | | |
| --- | --- | --- | --- | --- | --- | --- | --- | --- |
|  | **ssGSEA** | | | | **GSVA** | | | |
| **Signature** | **P value** | **Lower AUC 95%CI** | **AUC** | **Upper AUC 95% CI** | **P value** | **Lower AUC 95% CI** | **AUC** | **Upper AUC 95% CI** |
| **Anderson_42** | 0.3724 | 0.5121 | 0.5825 | 0.7294 | 0.971 | 0.5046 | 0.5175 | 0.674 |
| **Anderson_OD_51** | 0.0003 | 0.6397 | 0.8125 | 0.9279 | 0.0015 | 0.6139 | 0.7675 | 0.9013 |
| **Berry_393** | 0.0004 | 0.7183 | 0.83 | 0.9546 | 0.0018 | 0.6577 | 0.7875 | 0.9228 |
| **Berry_OD_86** | 0.0018 | 0.6029 | 0.77 | 0.9014 | 0.0018 | 0.5214 | 0.78 | 0.9067 |
| **Blankley_380** | 0.0003 | 0.6712 | 0.815 | 0.9282 | 0.0013 | 0.6075 | 0.7575 | 0.8782 |
| **Blankley_5** | 0.0001 | 0.6987 | 0.8275 | 0.9371 | 0.0017 | 0.6303 | 0.78 | 0.9161 |
| **Bloom_OD_144** | 0.0008 | 0.6712 | 0.785 | 0.9116 | 0.0071 | 0.651 | 0.77 | 0.909 |
| **Chen_5** | 0.0091 | 0.5854 | 0.7575 | 0.8946 | 0.0014 | 0.6123 | 0.78 | 0.9153 |
| **Chen_HIV_4** | 0.0033 | 0.6199 | 0.775 | 0.9106 | 0.0082 | 0.558 | 0.725 | 0.8946 |
| **Dawany_HIV_251** | 0.5773 | 0.5134 | 0.5625 | 0.7159 | 0.0503 | 0.517 | 0.695 | 0.8341 |
| **Duffy_23** | 0.5133 | 0.5019 | 0.5475 | 0.6876 | 0.3877 | 0.5066 | 0.59 | 0.7253 |
| **Esmail_203** | 0.0011 | 0.6498 | 0.79 | 0.9058 | 0.0601 | 0.5199 | 0.6825 | 0.8492 |
| **Esmail_82** | 0.0006 | 0.6399 | 0.7875 | 0.9266 | 0.0579 | 0.5317 | 0.6375 | 0.8495 |
| **Esmail_OD_893** | 0.0001 | 0.7199 | 0.8275 | 0.9485 | 0.0007 | 0.6681 | 0.785 | 0.9373 |
| **Estevez_133** | 0.0001 | 0.7178 | 0.8425 | 0.9398 | 0.0002 | 0.6489 | 0.81 | 0.9211 |
| **Estevez_259** | 0 | 0.7043 | 0.845 | 0.9497 | 0.0006 | 0.6521 | 0.795 | 0.9152 |
| **Francisco_OD_2** | 0.8679 | 0.5044 | 0.51 | 0.7019 | 0.0357 | 0.5246 | 0.695 | 0.8193 |
| **Gjoen_10** | 0.0171 | 0.5456 | 0.6975 | 0.8688 | 0.0825 | 0.52 | 0.665 | 0.8241 |
| **Gjoen_7** | 0.0009 | 0.5761 | 0.765 | 0.8883 | 0.0028 | 0.6067 | 0.77 | 0.8799 |
| **Gliddon_2_OD_4** | 0.0002 | 0.6953 | 0.835 | 0.95 | 0.0021 | 0.6001 | 0.77 | 0.8967 |
| **Gliddon_HIV_3** | 0 | 0.7274 | 0.8675 | 0.951 | 0.0073 | 0.5734 | 0.75 | 0.8815 |
| **Gliddon_OD_3** | 0 | 0.738 | 0.8675 | 0.9674 | 0.0073 | 0.6301 | 0.75 | 0.8942 |
| **Gliddon_OD_4** | 0.0002 | 0.6792 | 0.835 | 0.9299 | 0.0021 | 0.635 | 0.77 | 0.8812 |
| **Gong_OD_4** | 0.0002 | 0.6627 | 0.82 | 0.9502 | 0.0002 | 0.6597 | 0.815 | 0.9432 |
| **Hoang_OD_13** | 0.0002 | 0.7353 | 0.8625 | 0.9796 | 0.0015 | 0.6059 | 0.765 | 0.9127 |
| **Hoang_OD_20** | 0 | 0.7626 | 0.87 | 0.9786 | 0.0002 | 0.6868 | 0.825 | 0.9517 |
| **Huang_OD_13** | 0.0011 | 0.6147 | 0.795 | 0.9143 | 0.0124 | 0.5994 | 0.7325 | 0.872 |
| **Jacobsen_3** | 0.0001 | 0.7136 | 0.8325 | 0.9486 | 0.0037 | 0.655 | 0.78 | 0.9158 |
| **Jenum_8** | 0.0214 | 0.5368 | 0.69 | 0.8626 | 0.0448 | 0.5217 | 0.655 | 0.8309 |
| **Kaforou_27** | 0.0001 | 0.6455 | 0.85 | 0.97 | 0.0022 | 0.5998 | 0.78 | 0.9222 |
| **Kaforou_OD_44** | 0 | 0.7291 | 0.865 | 0.9676 | 0.0022 | 0.6116 | 0.7775 | 0.9184 |
| **Kaforou_OD_53** | 0.0002 | 0.6722 | 0.8375 | 0.9675 | 0.0088 | 0.5646 | 0.74 | 0.9305 |
| **Kaul_3** | 0.0036 | 0.6564 | 0.82 | 0.9223 | 0.0001 | 0.6872 | 0.8375 | 0.9498 |
| **Kulkarni_HIV_2** | 0.0016 | 0.6479 | 0.7725 | 0.9103 | 0.0012 | 0.6937 | 0.795 | 0.9239 |
| **Kwan_186** | 0.001 | 0.611 | 0.7725 | 0.8831 | 0.0056 | 0.5759 | 0.735 | 0.8634 |
| **LauxdaCosta_OD_3** | 0.0017 | 0.6111 | 0.77 | 0.8877 | 0.0027 | 0.609 | 0.7725 | 0.9 |
| **Lee_4** | 0.3147 | 0.5056 | 0.63 | 0.8015 | 0.8918 | 0.5075 | 0.525 | 0.7128 |
| **Leong_24** | 0.0035 | 0.5734 | 0.76 | 0.8858 | 0.2031 | 0.5125 | 0.6425 | 0.8046 |
| **Maertzdorf_15** | 0 | 0.7484 | 0.8825 | 0.976 | 0 | 0.7078 | 0.85 | 0.9652 |
| **Maertzdorf_4** | 0.0001 | 0.708 | 0.85 | 0.967 | 0.0096 | 0.5723 | 0.74 | 0.9139 |
| **Maertzdorf_OD_100** | 0.0866 | 0.5086 | 0.625 | 0.7678 | 0.0103 | 0.5969 | 0.7375 | 0.856 |
| **Natarajan_7** | 0.0019 | 0.6269 | 0.7575 | 0.9026 | 0.2395 | 0.5063 | 0.585 | 0.7754 |
| **Qian_OD_17** | 0.2342 | 0.5038 | 0.5775 | 0.7507 | 0.1653 | 0.5094 | 0.63 | 0.7617 |
| **Rajan_HIV_5** | 0 | 0.7644 | 0.8875 | 0.9849 | 0.0009 | 0.6203 | 0.7875 | 0.9086 |
| **Roe_3** | 0 | 0.699 | 0.855 | 0.9798 | 0 | 0.7458 | 0.87 | 0.9783 |
| **Roe_OD_4** | 0.034 | 0.513 | 0.67 | 0.8393 | 0.1369 | 0.5185 | 0.6275 | 0.7897 |
| **Sambarey_HIV_10** | 0.0007 | 0.6595 | 0.8075 | 0.9329 | 0.0012 | 0.643 | 0.79 | 0.9299 |
| **Singhania_OD_20** | 0.126 | 0.5216 | 0.65 | 0.8327 | 0.9561 | 0.5031 | 0.5025 | 0.6942 |
| **Sivakumaran_11** | 0.8887 | 0.5038 | 0.5 | 0.7151 | 0.5349 | 0.5086 | 0.5425 | 0.7248 |
| **Sloot_HIV_2** | 0.1956 | 0.5069 | 0.575 | 0.7487 | 0.3083 | 0.5113 | 0.5875 | 0.7366 |
| **Sweeney_OD_3** | 0 | 0.7539 | 0.8625 | 0.947 | 0.0024 | 0.6218 | 0.76 | 0.9093 |
| **Tabone_OD_11** | 0 | 0.769 | 0.885 | 0.9756 | 0 | 0.7612 | 0.8725 | 0.9855 |
| **Thompson_9** | 0 | 0.7071 | 0.855 | 0.96 | 0.0002 | 0.6726 | 0.83 | 0.9496 |
| **Tornheim_71** | 0 | 0.7547 | 0.8625 | 0.9562 | 0.0003 | 0.6717 | 0.8025 | 0.9233 |
| **Vargas_18** | 0.0001 | 0.712 | 0.8425 | 0.9683 | 0.0005 | 0.6447 | 0.805 | 0.9434 |
| **Vargas_42** | 0.0001 | 0.71 | 0.8425 | 0.9676 | 0.0006 | 0.6531 | 0.795 | 0.9335 |
| **Verhagen_10** | 0.6417 | 0.5038 | 0.5075 | 0.7236 | 0.5779 | 0.5013 | 0.5275 | 0.7376 |
| **Walter_51** | 0.0001 | 0.6858 | 0.8325 | 0.9375 | 0.0034 | 0.6226 | 0.7725 | 0.9094 |
| **Zhao_NANO_6** | 0.0715 | 0.51 | 0.69 | 0.8544 | 0.0511 | 0.5383 | 0.69 | 0.8082 |

*Supplementary Table 6: Results from the TBSignature Profiler evaluating the diagnostic performance of 59 published blood signatures in classifying TB status in the blood sequencing dataset. Two scoring methods were used: GSVA and ssGSEA. p-values were calculated using a 2-sample t-test. AUC values are shown with confidence intervals derived from bootstrapping. GSVA= gene set variation analysis; ssGSEA= single-sample GSEA, CI= confidence intervals.*
